# Racial and Ethnic Disparities in Years of Potential Life Lost Attributable to COVID-19 in the United States: An Analysis of 45 States and the District of Columbia

**DOI:** 10.1101/2021.01.28.21249411

**Authors:** Jay J. Xu, Jarvis T. Chen, Thomas R. Belin, Ronald S. Brookmeyer, Marc A. Suchard, Christina M. Ramirez

## Abstract

The coronavirus disease 2019 (COVID-19) epidemic in the United States has disproportionately impacted communities of color across the country. Focusing on COVID-19-attributable mortality, we expand upon a national comparative analysis of years of potential life lost (YPLL) attributable to COVID-19 by race/ethnicity (Bassett et al., 2020), estimating percentages of total YPLL for non-Hispanic Whites, non-Hispanic Blacks, Hispanics, non-Hispanic Asians, and non-Hispanic American Indian or Alaska Natives, contrasting them with their respective percent population shares, as well as age-adjusted YPLL rate ratios – anchoring comparisons to non-Hispanic Whites – in each of 45 states and the District of Columbia using data from the National Center for Health Statistics as of December 30, 2020. Using a novel Monte Carlo simulation procedure to quantify estimation uncertainty, our results reveal substantial racial/ethnic disparities in COVID-19-attributable YPLL across states, with a prevailing pattern of non-Hispanic Blacks and Hispanics experiencing disproportionately high and non-Hispanic Whites experiencing disproportionately low COVID-19-attributable YPLL. Furthermore, observed disparities are generally more pronounced when measuring mortality in terms of YPLL compared to death counts, reflecting the greater intensity of the disparities at younger ages. We also find substantial state-to-state variability in the magnitudes of the estimated racial/ethnic disparities, suggesting that they are driven in large part by social determinants of health whose degree of association with race/ethnicity varies by state.

## 1. Introduction

Racial and ethnic disparities in outcomes associated with coronavirus disease 2019 (COVID-19) in the United States have been the subject of widespread concern and societal discourse. Statistics on COVID-19 outcomes that local and state governmental health agencies in the United States (U.S.) publicly released did not initially include racial/ethnic demographic information on confirmed cases or deaths. Public pressure quickly grew for such information to be provided [1,2], with many local and state governmental health agencies quickly following suit. Racial/ethnic data on COVID-19 outcomes have revealed substantial racial/ethnic disparities, with non-Hispanic Blacks, Hispanics, and non-Hispanic American Indian or Alaska Natives experiencing disproportionately high numbers of cases, hospitalizations, and deaths [3]. Characterizing racial/ethnic disparities in COVID-19 outcomes is an active area of scientific research [4–23], and quantifying racial/ethnic disparities in the COVID-19 mortality burden has been a particular area of focus. For example, the Color of Coronavirus project of the American Public Media Research Lab [24] tracks COVID-19 death counts by race/ethnicity in each state, providing analyses and extended commentary on mortality rates by race/ethnicity and comparisons between the percentage of total deaths and the percent population share by race/ethnicity. Another similar data hub for COVID-19 mortality statistics by race/ethnicity is the COVID Racial Data Tracker of the COVID Tracking Project [25].

Given the substantially greater COVID-19 case fatality rates experienced by individuals in older age groups, COVID-19 death counts and mortality rates are predominantly determined by data from COVID-19 decedents in older age groups. Younger individuals, however, are also susceptible to death from COVID-19, which in principle represent greater unrealized years of life, economic productivity, and broader contributions to society compared to decedents of greater age. An alternative epidemiological measure of mortality that explicitly weights deaths that occur at earlier ages more heavily than deaths that occur at later ages is years of potential life lost (YPLL) [26]. YPLL for an individual fatality *i* is defined to be the difference between an upper reference age *𝒜* and age at death *a*_*i*_ if the difference is positive and zero otherwise:

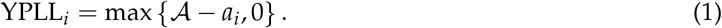

When comparing racial/ethnic subgroups with disparate mortality rates in younger age groups, higher YPLL is expected for those subgroups with comparatively higher mortality rates in the younger age groups, assuming a constant upper reference age is used across racial/ethnic groups. YPLL has been used in diverse contexts to quantify racial/ethnic disparities in premature mortality [27–29]. In the context of COVID-19, Bassett et al. (2020) [30] performed an analysis of COVID-19 deaths in the U.S. as a whole using COVID-19 mortality data from the National Center for Health Statistics as of July 22, 2020, calculating both age-specific mortality rates and YPLL by race/ethnicity. They found substantial racial/ethnic disparities in mortality rates across all age groups, but the magnitudes of the disparities were larger within younger age groups, which was reflected in their calculations of YPLL (with *𝒜* = 65). Although non-Hispanic Blacks and Hispanics both comprise a smaller percentage of the U.S. population than non-Hispanic Whites, Basset et al. found that both non-Hispanic Blacks and Hispanics experienced greater YPLL than non-Hispanic Whites, which was largely due to comparatively higher mortality rates in younger age groups.

Here, we extend the work of Bassett et al. by quantifying racial/ethnic disparities in YPLL attributable to COVID-19 at the individual state level, examining both their magnitude and their state-to-state variation. Specifically, we estimate percentages of total YPLL by race/ethnicity, contrasting them with their respective percent population shares, as well as age-adjusted YPLL rate ratios (RR) – anchoring comparisons to non-Hispanic Whites – in each of 45 states and the District of Columbia (D.C.). For comparison, we also calculate racial/ethnic percentages of total deaths and estimate the corresponding age-adjusted mortality RR’s to examine potential differences in the characterization of racial/ethnic disparities when measuring mortality in terms of YPLL compared to death counts. We also adopt a different inferential paradigm than Bassett et al. for uncertainty quantification, comprising novel Monte Carlo (MC) simulation techniques for interval estimation.

## 2. Materials and Methods

### 2.1. Data

We examine U.S. national COVID-19 mortality data from the National Center for Health Statistics (NCHS) as of December 30, 2020 – the last data update for calendar year 2020 – summarized as cumulative death counts within age groups stratified by state (as well as D.C. and Puerto Rico) and race/ethnicity [31], which we refer to hereafter as the NCHS Race/Ethnicity Data. The following 8 racial/ethnic groups are used in the NCHS Race/Ethnicity Data: non-Hispanic White (NH White), non-Hispanic Black (NH Black), Hispanic, non-Hispanic Asian (NH Asian), non-Hispanic American Indian or Alaska Native (NH AIAN), non-Hispanic Native Hawaiian or Other Pacific Islander, non-Hispanic Two or More Races, and Unknown. The set of mutually exclusive, collectively exhaustive, and chronologically ordered age groups used are <1, 1–4, 5–14, 15–24, 25–34, 35–44, 45–54, 55–64, 65–74, 75–84 and 85+. See File S1 in the Supplementary Materials for the NCHS Race/Ethnicity Data as of December 30, 2020, which consists of 301,679 total deaths. Because there is a lag in time between the actual date of death and when the death certificate is completed, submitted to NCHS, and processed, this number does not reflect the actual number of deaths in the U.S. as of December 30, 2020, which was reported to be 342,577 by the New York Times [32]. Some states’ data reported to the NCHS have been documented as severely delayed [33–35], especially North Carolina, which is one of a few states that, at the time of writing, doesn’t use an electronic death registration system [36]. Our analyses of the NCHS Race/Ethnicity Data implicitly assumes that there is no systematic bias in the speed with which death certificates are reported by states to the NCHS with respect to race/ethnicity or age at death.

Death counts between 1 and 9 within individual age groups are suppressed in the NCHS Race/Ethnicity Data due to confidentiality regulations. A total of 1,160 age groups across the 50 states plus D.C. and 8 racial/ethnic groups have suppressed death counts. Although the NCHS Race/Ethnicity Data do not explicitly provide the total number of actual deaths in each state, the NCHS provides a separate but synchronously updated publicly available dataset of COVID-19 deaths summarized as cumulative death counts within the same age groups as the NCHS Race/Ethnicity Data stratified by state and sex, which we refer to hereafter as the NCHS Sex Data [37], that does explicitly provide the total number of actual deaths in each state; see File S2 in the Supplementary Materials for the NCHS Sex Data as of December 30, 2020. Hence, for each state, we can ascertain the total number of deaths that are within the union of age groups with suppressed death counts, each of which contains between 1 and 9 deaths. The NCHS Race/Ethnicity Data also provides non-suppressed death counts within these same age groups stratified by race/ethnicity for the U.S. overall.

To standardize estimates by age, we make use of 2019 estimates of the U.S. population age distribution stratified by state, race, and ethnicity from CDC WONDER [38], defined over integer ages from 0 to 84 and a catch-all 85+ age group. Percent population shares by race/ethnicity both nationally and in each state are calculated from the 2019 CDC WONDER data. See File S3 in the Supplementary Materials for the 2019 CDC WONDER data.

### 2.2. Estimation Procedure for YPLL-Based Estimands from Administratively Interval Censored Ages at Death

The YPLL-based estimands that we consider here are total YPLL, percentage of total YPLL, and age-adjusted YPLL rates by race/ethnicity as well as age-adjusted YPLL rate ratios for NH Blacks, Hispanics, NH Asians, and NH AIAN’s relative to NH Whites. In contrast to the super-population perspective adopted by Basset et al., where observed COVID-19 deaths were viewed as a single realization of a hypothetical probability distribution and frequentist statistical theory was used for interval estimation [39], we take the finite-population perspective that the observed COVID-19 deaths simply constitutes the population of interest. As a result, estimation uncertainty in our context can be attributed to three sources: (a) administrative interval censoring of ages at death, (b) suppression of low death counts within individual age intervals, and (c) unknown race/ethnicity for a subset of deaths.

Focusing first on the issue of administrative interval censoring of ages at death and assuming no suppressed death counts or deaths of unknown race/ethnicity for purposes of illustration, the unknown exact ages at death for each individual precludes exact calculation of individual YPLL values. For the purpose of calculating aggregate YPLL, the standard approach in such settings is to operationally impute each individual’s age at death with its interval midpoint, a method that implicitly assumes uniformly distributed ages at death within individual age intervals [40]. However, applied epidemiological studies employing this “midpoint method” typically do not quantify the uncertainty associated with YPLL-based estimates as a result of the administrative interval censoring of ages at death [41–44]. Xu et al. (2021) [45] proposed a MC simulation procedure to quantify the uncertainty associated with YPLL-based estimates obtained from mortality data summarized as death counts within age intervals. We refer the reader to their paper for the details of the MC simulation procedure, but to summarize it briefly, the procedure comprises independently simulating ages at death for each individual from continuous uniform distributions defined over their respective age intervals at each MC iteration, calculating a point estimate of the estimand of interest from the corresponding simulated YPLL values. The overall point estimate is defined to be the mean of the collection of MC point estimates, and the lower and upper endpoints of a (1 − *α*) × 100% interval estimate (which can be conceptualized as a “range interval” per Bobashev and Morris (2010) [46]) are defined to be the 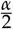 and 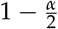 quantiles of the collection of MC point estimates, respectively.

### 2.3. Procedure Modification to Account for Suppression of Low Death Counts

The second source of estimation uncertainty, suppression of low death counts within individual age intervals, arises out of privacy and confidentiality concerns associated with publishing small death counts within individual age intervals, which can risk revealing personally identifiable information for individual COVID-19 deaths. We introduce a modification of the Xu et al. MC simulation procedure described in Section 2.2 to account for this additional source of estimation uncertainty while still assuming no deaths of unknown race/ethnicity for purposes of illustration.

For each state, we know the total number of deaths contained in the union of intervals with suppressed death counts, each of which must be an integer between 1 and 9. As such, we can exhaustively enumerate all possible death count combinations across the intervals with suppressed death counts. Each death count combination corresponding to the intervals with suppressed death counts juxtaposed with the intervals containing non-suppressed death counts constitutes one possible “mortality dataset” of death counts within age intervals. We modify the Xu et al. MC simulation procedure by independently simulating ages at death for each individual for each mortality dataset at each MC iteration. Then, a point estimate of the estimand of interest is calculated for each mortality dataset from the corresponding simulated YPLL values, from which we store only the minimum and maximum point estimate across the mortality datasets at each MC iteration. A conservative (1 − *α*) × 100% interval estimate of the estimand of interest is then constructed from the 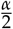 quantile of the collection of minimum MC point estimates and the 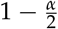 quantile of the collection of maximum MC point estimates. We describe the interval estimate as “conservative” as a result of our estimation strategy of considering all possible mortality datasets and using the extrema of the subsequent MC point estimates to construct the interval estimate; a (1 − *α*) × 100% interval estimate of the estimand of interest obtained from the standard Xu et al. MC simulation procedure had the suppressed death counts been known (and assuming no deaths with unknown race/ethnicity) would be completely contained in the corresponding conservative (1 − *α*) × 100% interval estimate.

### 2.4. Procedure Modification to Further Account for a Subset of Deaths with Unknown Race/Ethnicty

The third source of estimation uncertainty, unknown race/ethnicity for a subset of deaths, can be accommodated through an additional modification of the Xu et al. MC simulation procedure. The key idea is to separate the tasks of obtaining a maximum and minimum point estimate at each MC iteration. Specifically, when the estimand of interest primarily concerns racial/ethnic group *r* (i.e., total YPLL for racial/ethnic group *r*, percentage of total YPLL for racial/ethnic group *r*, age-adjusted YPLL rate for racial/ethnic group *r*, and age-adjusted *r*-to-NH White YPLL RR), we assume all deaths among the Unknown racial/ethnic group are members of racial/ethnic group *r* and appropriately combine death counts within their age intervals before exhaustively enumerating all possible mortality datasets, simulating ages at death for each individual for each mortality dataset, calculating a point estimate from the corresponding simulated YPLL values for each mortality datset, and storing the maximum point estimate across mortality datasets at each MC iteration. The task of obtaining a minimum point estimate at each MC iteration is pursued analogously but separately, where the key distinction is that we do not combine the Unknown racial/ethnic group and racial/ethnic group *r*. Therefore, a separate set of mortality datasets is considered for each task of maximization and minimization.

### 2.5. Computational Savings by Omitting Unnecessary Mortality Datasets

The total number of mortality dataset scenarios can be enormous, potentially making it computationally infeasible to simulate ages at death for all mortality datasets. However, substantial computational savings can be achieved by (a) combining racial/ethnic groups not included in the definition of the estimand of interest and (b) identifying mortality datasets that we do not need to simulate ages at death from because they would yield a maximum or minimum MC point estimate with probability 0. We describe an example of such combinatorial reductions when the estimand of interest is the percentage of total YPLL for racial/ethnic group *r*.

For the task of obtaining a maximum point estimate at each MC iteration, we combine the Unknown racial/ethnic group with racial/ethnic group *r*, combine the remaining racial/ethnic groups into a single non-*r* racial/ethnic group, which we denote 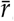, and enumerate all possible mortality datasets. In this case, only one mortality dataset needs to be considered, namely, the one that contains the maximum possible number of deaths in the lowest age intervals corresponding to suppressed death counts for racial/ethnic group *r*. The remaining mortality datasets can be omitted because they would yield a maximum MC point estimate with probability 0. For the task of obtaining a minimum point estimate at each MC iteration, we combine all of the non-*r* racial/ethnic groups (including the Unknown racial/ethnic group) into a single non-*r* racial/ethnic group 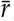 and enumerate all possible mortality datasets. Here, the only mortality dataset that needs to be considered is the one that contains the maximum possible number of deaths in the lowest age intervals corresponding to suppressed death counts for the combined non-*r* racial/ethnic group 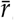 and, if necessary, the maximum possible number of deaths in the highest age intervals corresponding to suppressed death counts for racial/ethnic group *r*. The remaining mortality datasets can be omitted because they would yield a minimum MC point estimate with probability 0.

For estimation of the age-adjusted YPLL rates and RR’s, identifying mortality datasets that we not need to simulate ages at death from is a more challenging task, but it is still possible. Inequality conditions can be established computationally to identify meaningful numbers of mortality datasets that can be omitted in our analyses.

### 2.6. Complete Monte Carlo Simulation Procedure for Estimation of YPLL-Based Estimands

Here, we comprehensively summarize the complete modified Xu et al. MC simulation procedure for estimation of the YPLL-based estimands of interest that quantifies the uncertainty associated with the administrative interval censoring of ages at death, suppression of low death counts within individual age intervals, and unknown race/ethnicity for a subset of deaths. We limit the analyses presented here to jurisdictions in the continental U.S. with at least 500 total deaths in the NCHS Sex Data, a condition satisfied by 45 states as well as D.C., which for brevity we characterize as a “state” in summaries of the results of our analyses. The 5 states omitted in our analyses are Alaska, Hawaii, Maine, Vermont, and Wyoming. For each examined state *s*, the procedure can be comprehensively summarized as follows.

1. Calculate the difference between the total number of deaths as provided by the NCHS Sex Data and the total number of deaths contained in intervals with non-suppressed death counts; this is the number of deaths contained in the union of intervals with suppressed death counts.
2. Let *ℬ* denote the total number of MC iterations, and let *b* = 1, …, *ℬ* index the MC iterations. For the task of obtaining a maximum MC point estimate of the estimand of interest that primarily concerns racial/ethnic group *r* at each MC iteration *b*, combine the Unknown racial/ethnic group with racial/ethnic group *r* into a combined *r*-Unknown racial/ethnic group whose constituents we all assume to be in racial/ethnic group *r*, combine the remaining racial/ethnic groups that are not included in the definition of the estimand into a single “other” racial/ethnic group, and enumerate all possible mortality datasets, omitting those that would yield a maximum MC point estimate with probability 0. For the task of obtaining a minimum MC point estimate of the estimand of interest, combine all of the racial/ethnic groups not included in the definition of the estimand (including the Unknown racial/ethnic group) and enumerate all possible mortality datasets, omitting those that would yield a minimum MC point estimate with probability 0. Let 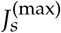 denote the number of mortality datasets considered for the maximization task in state *s*, and let 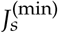 denote the number of mortality datasets considered for the minimization task in state *s*.
3. Specify a YPLL upper reference age *𝒜* less than or equal to 85 years. We view age group <1 as equivalent to the singular age 0, and the remaining numeric NCHS age group endpoints represent integer age at last birthday so that there is a 1-year gap between the endpoints of two chronologically consecutive age groups (e.g., 35-44 and 45-54). We treat age as a continuous variable, and as a consequence, we mathematically interpret the <1 age group (age 0) as the right half-open interval [0, 1), the 85+ age group as the half-bounded interval [85, ∞), and the remaining NCHS age groups as right half-open intervals with lower limit equal to the lower endpoint of the corresponding NCHS age group and upper limit equal to the upper endpoint of the corresponding NCHS age group plus one (e.g., age group 5–14 is viewed as = [5, 15)). At each MC iteration *b* and for each mortality dataset 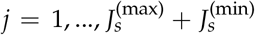 considered for the maximization and minimization tasks, independently simulate an age at death 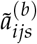 for each individual fatality *i* corresponding to age interval (*L*_*ijs*_, *U*_*ijs*_), where *𝒜* > *L*_*ijs*_, from the corresponding continuous uniform distribution:

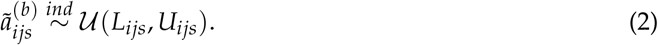 Observe that *𝒜* is intentionally and necessarily chosen to be less than or equal to 85 years to obviate the simulation of ages at death corresponding to the 85+ age group, thereby avoiding potential analytic difficulties because each fatality corresponding to the 85+ age group contributes nothing to YPLL.
4. At each MC iteration *b* and for each mortality dataset *j*, calculate a point estimate of the estimand of interest. In particular, for the estimation of the percentage of total YPLL for racial/ethnic group *r*, first calculate total YPLL for racial/ethnic group *r* (which includes the Unknown racial/ethnic group for the maximization task) and the remaining racial/ethnic groups from the simulated ages at death, which are 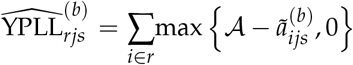 and 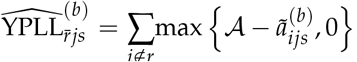, respectively. Then, the percentage of total YPLL for racial/ethnic group *r*, which we denote 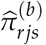 is given by:

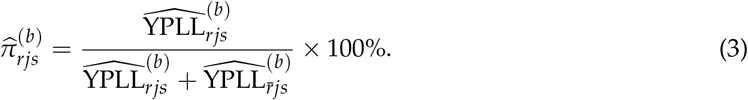 For estimation of the age-adjusted *r*-to-NH White YPLL RR, first estimate the age-adjusted YPLL rates for racial/ethnic group *r* and NH Whites, using the 2019 CDC WONDER age distribution estimate of the overall U.S. population as the standard population. The age-adjusted YPLL rate for racial/ethnic group *r* (which includes the Unknown racial/ethnic group for the maximization task) is calculated using direct age adjustment [47] from the simulated ages at death, which we denote 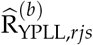. Since the simulated ages at death are continuous and the CDC WONDER age distribution estimates are defined over integer ages from 0 to 84, we aggregate the corresponding simulated YPLL values with respect the 1-year intervals implied by these integer ages (i.e., age *a* ∈ {0, 1, …, 84} implies age interval [*a, a* + 1)) to calculate the age-specific YPLL rates, which are subsequently applied to the standard population to obtain the age-adjusted YPLL rate for racial/ethnic group *r*:

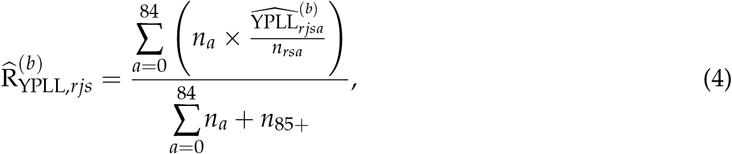

where 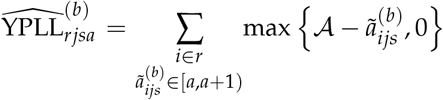 denotes aggregate YPLL corresponding to age *a* ∈ {0, 1, …, 84}; *n*_*rsa*_ denotes the 2019 CDC WONDER U.S. population estimate for race/ethnicity *r*, state *s*, and age *a* ∈ {0, 1, …, 84}; and *n*_*a*_ denotes the 2019 CDC WONDER U.S. population estimate for age *a* ∈ {0, 1, …, 84, 85+}. Then, the age-adjusted *r*-to-NH White YPLL RR is defined to be the quotient of 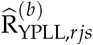 and 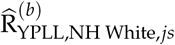:

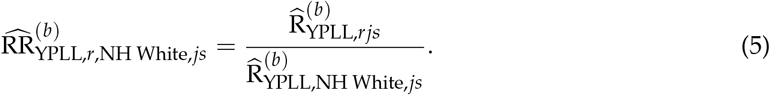
5. At each MC iteration *b*, store the maximum of the 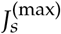 MC point estimates of the estimand of interest calculated from the set of 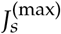 mortality datasets considered for the maximization task, and store the minimum of the 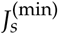 MC point estimates calculated from the set of 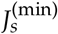mortality datasets considered for the minimization task.
6. A conservative (1 − *α*) × 100% interval estimate of the estimand of interest is given by the 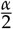 quantile of the *ℬ* minimum MC point estimates and the 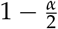.quantile of the *ℬ* maximum MC point estimates.

Defining a valid overall point estimate for the estimand of interest is not straightforward due to our estimation strategy; to be explicit, the midpoint of the conservative (1 − *α*) × 100% interval estimate should not be interpreted as the point estimate. As such, in the results we present, we communicate our estimates solely as intervals, basing conclusions and interpretations on the ensemble of interval estimates we produce.

### 2.7. Monte Carlo Simulation Procedure for Estimation of Age-Adjusted Mortality Rates and Rate Ratios

Because we can enumerate the death count combinations for the intervals with suppressed death counts in each state, we can determine the entire plausible range of total deaths and percentage of total deaths by race/ethnicity in each state, which complement our estimates of total YPLL and percentage of total YPLL by race/ethnicity. At the time of writing, every U.S. state except North Dakota publicly reports COVID-19 death counts by race/ethnicity [48], which are more up to date than the (lagged) NCHS Race/Ethnicity Data. However, because different states can use different race/ethnicity categories, rendering direct comparisons between states difficult, we did not use the COVID-19 death counts by race/ethnicity that states themselves publicly report in our analyses. For comparison to our estimates of the age-adjusted YPLL rates and RR’s, we also consider estimation of the corresponding age-adjusted mortality rates and RR’s. We want these quantities to be age-standardized to the 2019 CDC WONDER age distribution estimate of the overall U.S. population – without combining CDC WONDER age intervals to align them with the NCHS age intervals – so that estimated YPLL and mortality rates in our analyses are age-standardized to as identical as possible standard populations in terms of age interval granularity. To this end, we perform an analogous MC simulation procedure to obtain conservative (1 − *α*) × 100% interval estimates of the age-adjusted mortality rates and RR’s in the U.S. and in each examined state. The procedure largely mirrors the MC simulation procedure for the YPLL-based estimands described previously in Section 2.6. However, ages at death are simulated for all individuals in non-85+ age groups. In the direct age adjustment procedure, we sum the number of simulated ages at death falling within the 1-year intervals implied by integer ages 0 to 84 to calculate the age-specific mortality rates for ages 0 to 84 as well as calculate an age 85+ mortality rate, which are subsequently applied to the standard population to obtain the age-adjusted mortality rate, which we denote 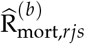 for racial/ethnic group *r*:

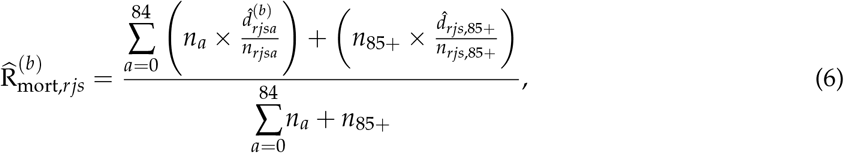

where 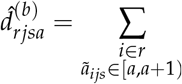 1 is the number of simulated ages at death equal to age *a* ∈ {0, 1, …, 84}, and 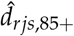 denotes the number of deaths in the 85+ age group. Then, the age-adjusted *r*-to-NH White mortality RR is defined to be the quotient of 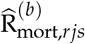 and 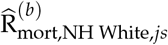:

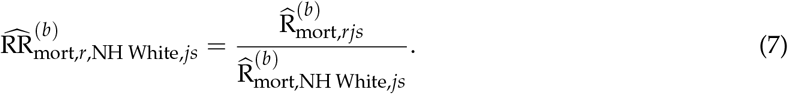

### 2.8. Computation

We perform the modified Xu et al. MC simulation procedure described in Section 2.6 for *ℬ* = 1000 iterations using a constant YPLL upper reference age of *𝒜* = 75 years for all racial/ethnic groups considered, an approach used by the Centers for Disease Control and Prevention (CDC) [49–56] and in other applied research domains [57,58], obtaining conservative 95% interval estimates of total YPLL, percentage of total YPLL, and age-adjusted YPLL rates by race/ethnicity as well as age-adjusted YPLL RR’s for NH Blacks, Hispanics, NH Asians, and NH AIAN’s relative to NH Whites in each of the 46 examined states. An approach that has been employed in applied studies contrasting YPLL by race/ethnicity is to use race/ethnicity-specific YPLL upper reference ages corresponding to the respective life expectancies [28,29,59–62], thereby reflecting known heterogeneity in underlying life expectancies between racial/ethnic groups. However, out of a desire informed by a health equity perspective not to necessarily normalize underlying racial/ethnic disparities in life expectancy, a sentiment shared in other applications [63,64], we decided to use a constant YPLL upper reference age (*𝒜* = 75) for all racial/ethnic groups considered to provide a comparison of YPLL by race/ethnicity that is not dependent on underlying racial/ethnic differences in life expectancies. We also obtain conservative 95% interval estimates of age-adjusted mortality rates by race/ethnicity as well as age-adjusted mortality RR’s for NH Blacks, Hispanics, NH Asians, and NH AIAN’s relative to NH Whites using an analogous MC simulation procedure described in Section 2.7, also for *ℬ* = 1000 iterations. All MC simulation analyses were performed using the R version 3.6.0 programming language [65]. The code used in the analyses is available upon reasonable request from the corresponding author.

## 3. Results

Tables S1-S5 in the Supplementary Materials contain the entirety of the results. Table S1 presents conservative 95% interval estimates of total YPLL for the 5 racial/ethnic groups considered in the U.S. and in each of the 46 examined states, and Table S2 presents intervals denoting the entire plausible range of total deaths by race/ethnicity in the U.S. and in each examined state. Table S3 presents conservative 95% interval estimates of the percentage of total YPLL and intervals denoting the entire plausible range of the percentage of total deaths for the 5 racial/ethnic groups considered in the U.S. and in each of the examined states. When the interval of the percentage of total deaths for racial/ethnic group *r* ∈ {NH White, NH Black, Hispanic, NH Asian, NH AIAN} is completely above its percent population share, deaths among racial/ethnic group *r* are overrepresented in COVID-19 deaths. Conversely, when the interval of the percentage of total deaths for racial/ethnic group *r* is completely below its percent population share, deaths among racial/ethnic group *r* are underrepresented in COVID-19 deaths. The interpretation of the conservative 95% interval estimates of the percentage of total YPLL relative to the percent population share is slightly more nuanced, however. When the conservative 95% interval estimate of the percentage of total YPLL for racial/ethnic group *r* is completely above its percent population share, deaths among racial/ethnic group *r* are either overrepresented or decedent ages among racial/ethnic group *r* are disproportionately younger relative to decedents of the other racial/ethnic groups or both – to a degree that is statistically discernable. Conversely, when the conservative 95% interval estimate of the percentage of total YPLL for racial/ethnic group *r* is completely below its percent population share, deaths among racial/ethnic group *r* are either underrepresented or decedent ages among racial/ethnic group *r* are disproportionately older relative to decedents of the other racial/ethnic groups or both – to a degree that is statistically discernable.

The magnitudes of the disparities can depend on whether mortality is measured in terms of YPLL or death counts. If the interval of the percentage of total deaths for racial/ethnic group *r* is completely above its percent population share and the conservative 95% interval estimate of the percentage of total YPLL for racial/ethnic group *r* is completely above the interval of the percentage of total deaths for racial/ethnic group *r*, it means that although deaths among racial/ethnic group *r* are overrepresented, decedent ages among racial/ethnic group *r* are also skewed disproportionately toward younger ages to such a degree that the disparity in the COVID-19 mortality burden with respect to racial/ethnic group *r* is statistically discernably greater when measuring mortality in terms of YPLL compared to death counts. Similarly, if the interval of the percentage of total deaths for racial/ethnic group *r* is completely below its percent population share and the conservative 95% interval estimate of the percentage of total YPLL for racial/ethnic group *r* is completely below the interval of the percentage of total deaths for racial/ethnic group *r*, it means that although deaths among racial/ethnic group *r* are underrepresented, decedent ages among racial/ethnic group *r* are also skewed disproportionately toward older ages to such a degree that the disparity in the COVID-19 mortality burden with respect to racial/ethnic group *r* is statistically discernably greater when measuring mortality in terms of YPLL compared to death counts.

Moreover, the direction of the disparities in the COVID-19 mortality burden can also depend on whether mortality is measured in terms of YPLL or death counts. If the interval of the percentage of total deaths for racial/ethnic group *r* is completely below its percent population share but the conservative 95% interval estimate of the percentage of total YPLL for racial/ethnic group *r* is completely above its percent population share, deaths among racial/ethnic group *r* are underrepresented, but decedent ages among racial/ethnic group *r* are skewed disproportionately toward younger ages relative to decedents of the other racial/ethnic groups to a degree that outweighs the disproportionately low number of deaths. Similarly, if the interval of the percentage of total deaths for racial/ethnic group *r* is completely above its percent population share but the conservative 95% interval estimate of the percentage of total YPLL for racial/ethnic group *r* is completely below its percent population share, deaths among racial/ethnic group *r* are overrepresented, but decedent ages among racial/ethnic group *r* are skewed disproportionately toward older ages relative to decedents of the other racial/ethnic groups to a degree that outweighs the disproportionately high number of deaths.

Table S4 presents conservative 95% interval estimates of the age-adjusted YPLL and mortality rates per 10,000 population for the 5 racial/ethnic groups considered in the U.S. and in the examined states. Table S5 presents conservative 95% interval estimates of the age-adjusted YPLL and mortality RR’s for NH Blacks, Hispanics, NH Asians, and NH AIAN’s relative to NH Whites in the U.S. and in the 46 examined states. When the age-adjusted *r*-to-NH White YPLL RR (*r* ∈ {NH Black, Hispanic, NH Asian, NH AIAN}) is completely above 1.0, it means that after accounting for differences in the population age distributions between racial/ethnic group *r* and NH Whites, racial/ethnic group *r* experiences COVID-19-attributable YPLL at a rate that that is statistically discernably above that of NH Whites. Similarly, when the age-adjusted *r*-to-NH White mortality RR is completely above 1.0, it means that after accounting for differences in the population age distributions between racial/ethnic group *r* and NH Whites, racial/ethnic group *r* experiences death from COVID-19 at a rate that is statistically discernably above that of NH Whites. When the conservative 95% interval estimate of the age-adjusted *r*-to-NH White YPLL RR is completely above the conservative 95% interval estimate of the age-adjusted *r*-to-NH White mortality RR, the age-adjusted COVID-19 mortality burden for racial/ethnic group *r* relative to NH Whites is statistically discernably greater in magnitude when measuring mortality in terms of YPLL compared to death counts as a result of individuals of racial/ethnic group *r* dying at systematically and statistically discernably younger ages relative to NH Whites after accounting for differences in the population age distributions between racial/ethnic group *r* and NH Whites.

### 3.1. Results for Non-Hispanic Whites

Figure 1 displays a graphical comparison between the conservative 95% interval estimates of the percentage of total YPLL, intervals denoting the entire plausible range of the percentage of total deaths, and the percent population share for NH Whites in the U.S. and in each examined state. Nationally, NH White COVID-19 deaths are underrepresented, comprising 57.6%–58.1% of total deaths despite representing 61.2% of the U.S. population. At the individual state level, NH White intervals of the percentage of total deaths are completely above the NH White percent population share in 24 of the examined states and completely below in 21 of the examined states. However, NH White COVID-19 decedent ages in the U.S. overall are skewed disproportionately toward older ages to such a degree that the U.S. national NH White conservative 95% interval estimate of the percentage of total YPLL ([40.1% 40.8%]) is completely below the U.S. national NH White interval of the percentage of total deaths. At the individual state level, the NH White conservative 95% interval estimates of the percentage of total YPLL are completely below the NH White intervals of the percentage of total deaths in 44 of the 46 examined states, reflecting a consistent pattern across states of NH Whites dying from COVID-19 at older ages than individuals of other racial/ethnic groups. For example, in Louisiana, NH Whites represent 59.1% of the population and only 52.6%–53.4% of total deaths, but the NH White conservative 95% interval estimate of the percentage of total YPLL is an even lower [29.7% – 34.7%]. Furthermore, NH White conservative 95% interval estimates of the percentage of total YPLL are completely below the NH White percent population share in 43 of the 46 examined states, greater than the corresponding number for the intervals of the percentage of total deaths. The direction of the disparity in fact reverses when using YPLL rather than death counts to measure mortality in 21 of the 24 states where NH White COVID-19 deaths are overrepresented. For example, in Massachusetts, NH Whites represent 72.1% of the population and 80.1%–81.3% of total deaths, but the NH White conservative 95% interval estimate of the percentage of total YPLL is only [54.0% – 62.7%].

**Figure 1.**
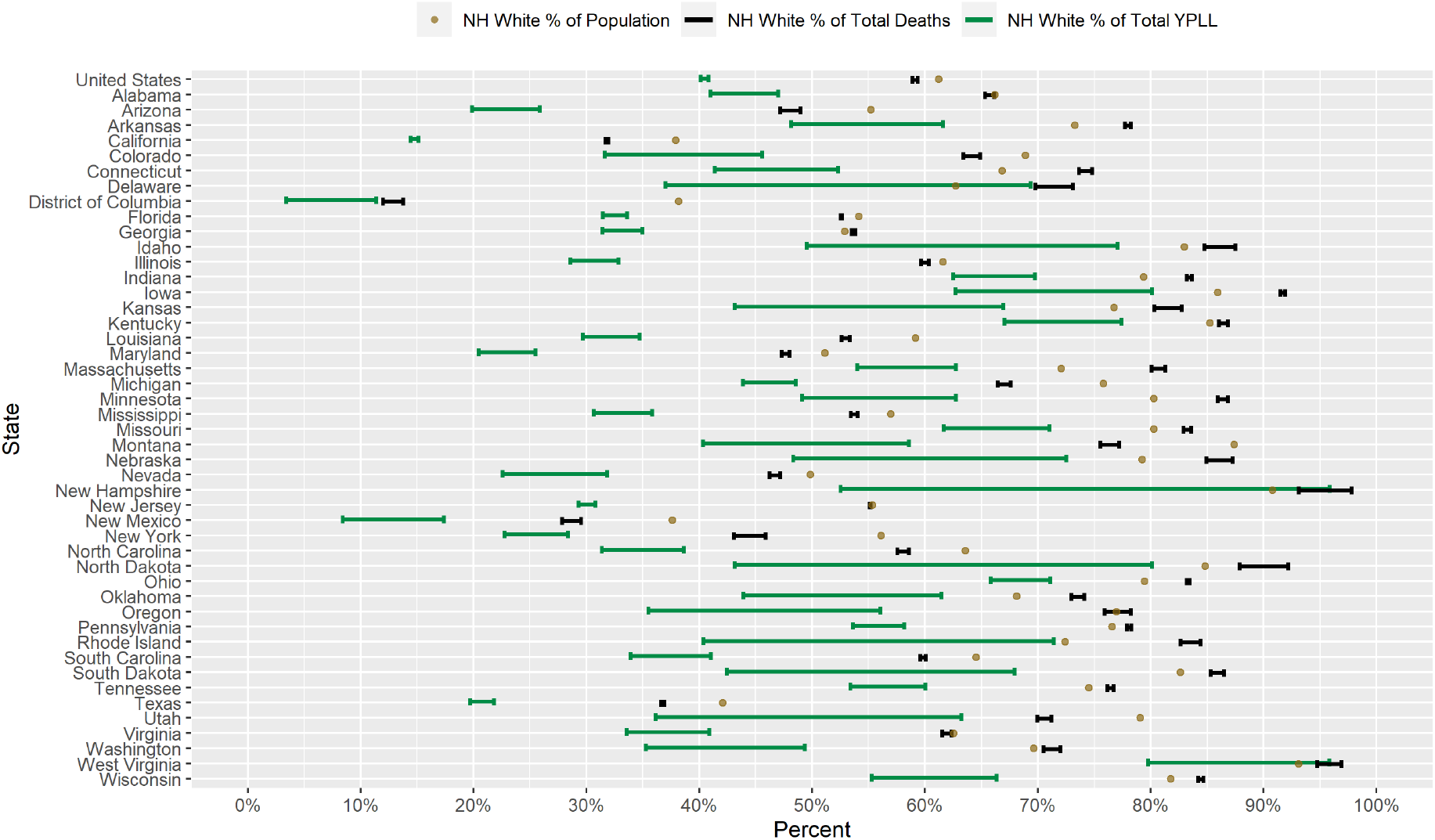
Conservative 95% interval estimates of the percentage of total YPLL before age 75, intervals denoting the entire plausible range for the percentage of total deaths, and the percent population shares for NH Whites in the U.S. and in each of the 45 examined states and the District of Columbia with respect to cumulative COVID-19 deaths according to data from the National Center for Health Statistics as of December 30, 2020.

### 3.2. Results for Non-Hispanic Blacks

Figure 2 displays a graphical comparison between the conservative 95% interval estimates of the percentage of total YPLL, intervals denoting the entire plausible range of the percentage of total deaths, and the percent population share for NH Blacks in the U.S. and in each examined state. Nationally, COVID-19 deaths among NH Blacks are overrepresented, comprising 16.9%–17.4% of total deaths despite representing only 13.2% of the U.S. population. Indeed, at the individual state level, NH Black intervals of the percentage of total deaths are completely above the NH Black percent population share in 26 of the examined states and completely below in only 8 of the examined states. In fact, the NH Black interval of the percentage of total deaths in D.C. is completely above all of the corresponding intervals for the other racial/ethnic groups. Although this is partially due to the fact that NH Blacks are the predominant racial/ethnic group in D.C., comprising over 45% of the population, the disparity between the percentage of total deaths and the NH Black percent population share is by far the highest in D.C. among the examined states. Moreover, NH Black COVID-19 decedent ages in the U.S. overall are skewed disproportionately toward younger ages to such a degree that the U.S. national NH Black conservative 95% interval estimate of the percentage of total YPLL ([23.0% – 23.6%]) is completely above the U.S. national NH Black interval of the percentage of total deaths. Indeed, at the individual state level, the NH Black conservative 95% interval estimates of the percentage of total YPLL are completely above the NH Black intervals of the percentage of total deaths in 27 of the examined states and completely below in none of the examined states. For example, NH Blacks represent 14.6% of the population in Michigan but an astonishing 27.7%–29.0% of total deaths, yet the NH Black conservative 95% interval estimate of the percentage of total YPLL is a staggering [42.4% – 47.4%]. Interestingly, the reverse phenomenon is observed in D.C., where the NH Black conservative 95% interval estimate of the percentage of total YPLL ([52.3% – 68.9%]) is completely below the NH Black interval of the percentage of total deaths ([69.6% – 71.4%]). NH Black conservative 95% interval estimates of the percentage of total YPLL are completely above the NH Black percent population share in 33 of the 46 examined states, greater than the corresponding number for the intervals of the percentage of total deaths, and completely below in none of the examined states. NH Black conservative 95% interval estimates of the percentage of total YPLL are also completely above all of the corresponding intervals for the other racial/ethnic groups in 6 of the examined states (D.C., Georgia, Louisiana, Maryland, Mississippi, and South Carolina). The direction of the disparity in fact reverses when using YPLL rather than death counts to measure mortality in 3 of the 8 states (Arkansas, Iowa, and Minnesota) where the NH Black intervals of the percentage of total deaths are completely below the NH Black percent population share. For example, in Minnesota, NH Blacks represent 7.5% of the population and only 5.3%–6.2% of total deaths, but the NH Black 95% conservative interval estimate of the percentage of total YPLL is [12.9% – 21.7%].

**Figure 2.**
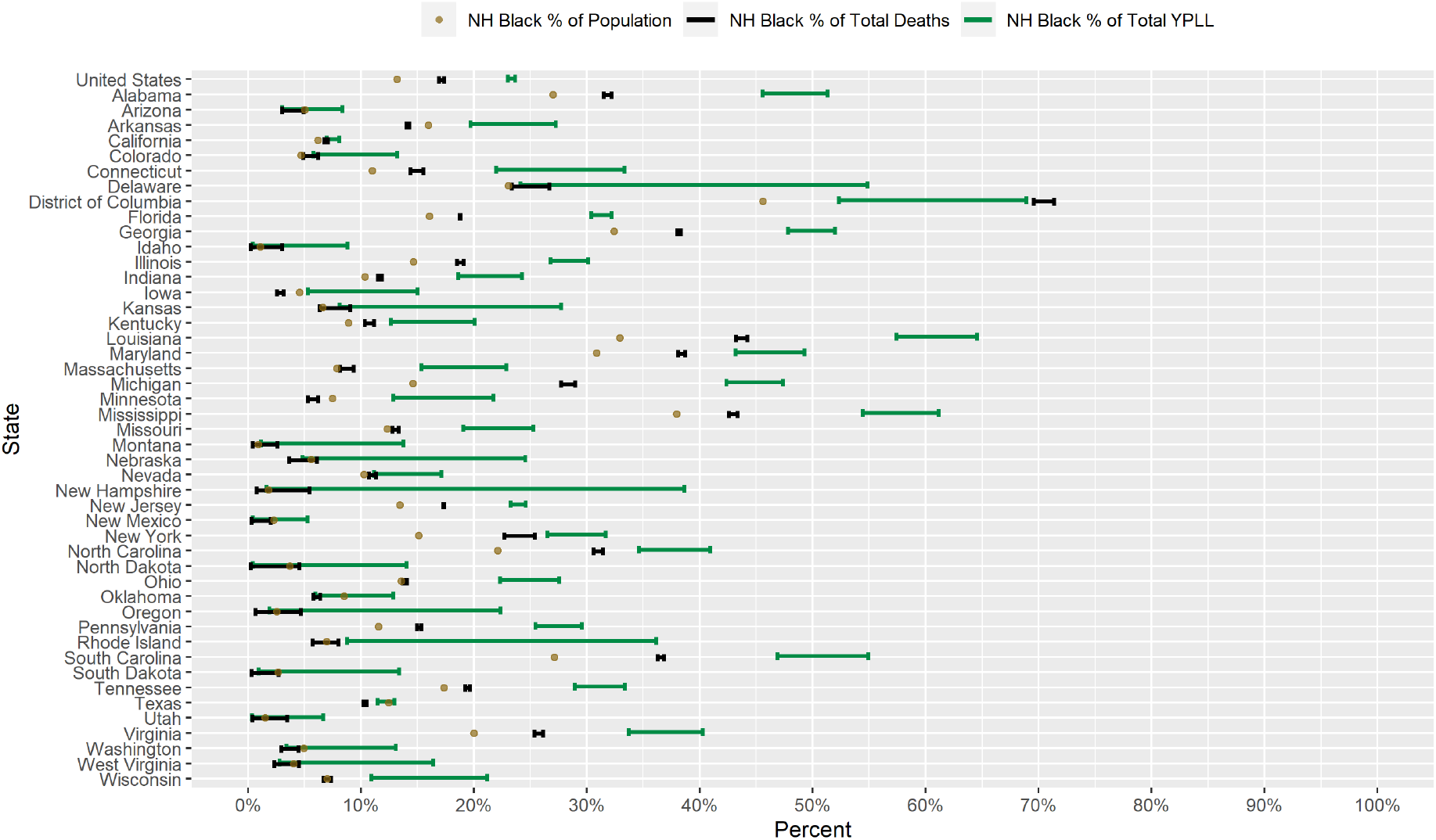
Conservative 95% interval estimates of the percentage of total YPLL before age 75, intervals denoting the entire plausible range for the percentage of total deaths, and the percent population shares for NH Blacks in the U.S. and in each of the 45 examined states and the District of Columbia with respect to cumulative COVID-19 deaths according to data from the National Center for Health Statistics as of December 30, 2020.

Figure 3 presents a graphical comparison of the conservative 95% interval estimates of the age-adjusted NH Black-to-NH White YPLL and mortality RR’s in the U.S. and in each examined state. The U.S. national conservative 95% interval estimate of the age-adjusted NH Black-to-NH White mortality RR is [1.91 – 1.99], and the conservative 95% interval estimates of the age-adjusted NH Black-to-NH White mortality RR are completely above 1.0, 2.0, and 3.0 in 39, 17, and 2 of the examined states, respectively. However, NH Black decedent ages in the U.S. overall are skewed disproportionately toward younger ages relative to NH Whites, after accounting for differences in the national population age distributions between NH Blacks and NH Whites, to such a degree that the U.S. national conservative 95% interval estimate of the age-adjusted NH Black-to-NH White YPLL RR is [2.84 – 2.97], completely above the U.S. national conservative 95% interval estimate of the age-adjusted NH Black-to-NH White mortality RR. Indeed, in 30 of the 46 examined states, the conservative 95% interval estimates of the age-adjusted NH Black-to-NH White YPLL RR are completely above the conservative 95% interval estimates of the age-adjusted NH Black-to-NH White mortality RR, reflecting a broad pattern across states of NH Blacks dying from COVID-19 at earlier ages than NH Whites after accounting for differences in their respective state population age distributions. Moreover, conservative 95% interval estimates of the age-adjusted NH Black-to-NH White YPLL RR are completely above 1.0, 2.0 and 3.0 in 39, 33, and 17 of the examined states, respectively, greater than or equal to the corresponding numbers for the age-adjusted NH Black-to-NH White mortality RR’s. Among the examined states, Michigan stands out markedly with the highest conservative 95% interval estimate of the age-adjusted NH Black-to-NH White YPLL RR ([5.77 – 6.67]).

**Figure 3.**
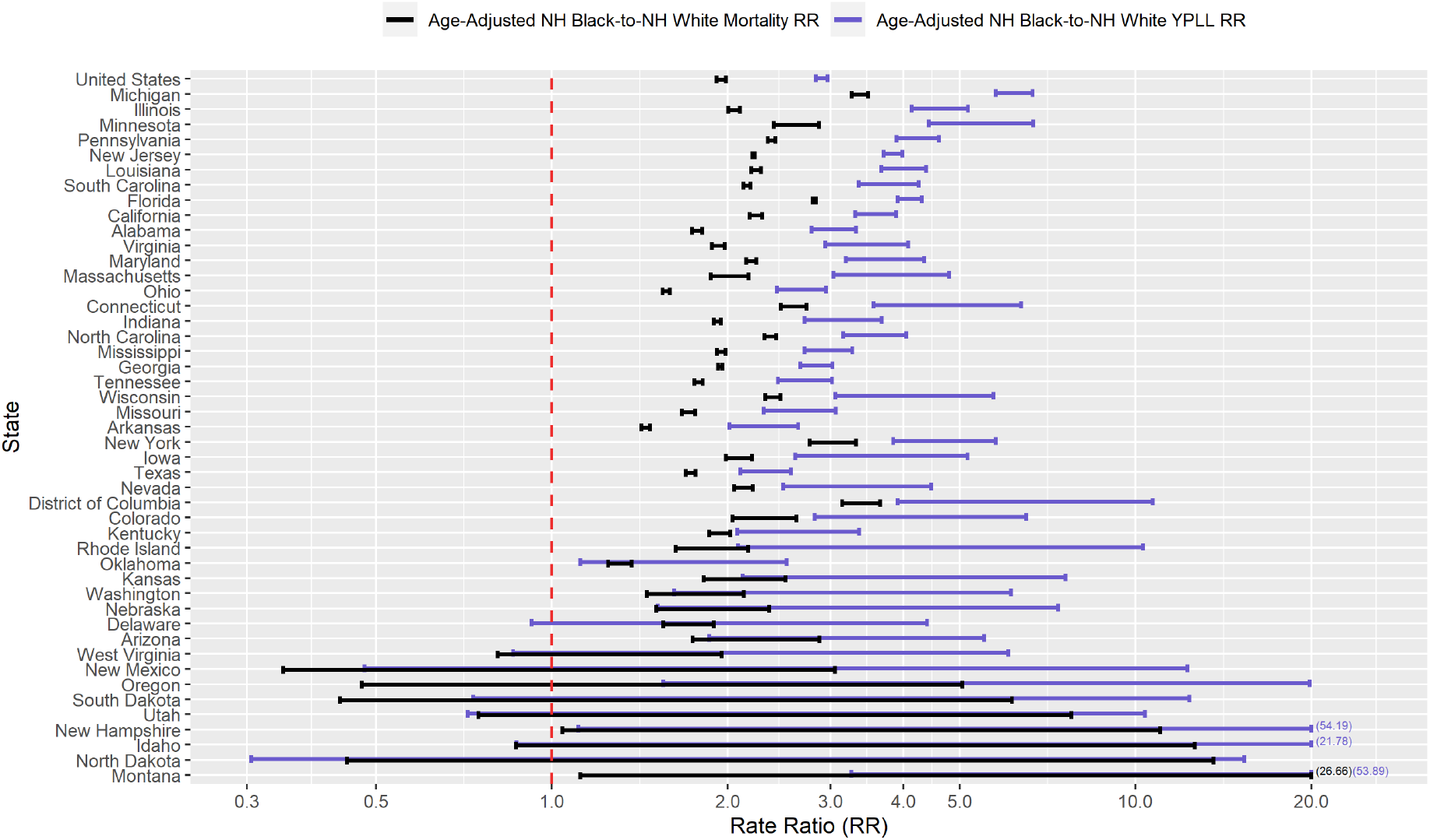
Conservative 95% interval estimates of the age-adjusted NH Black-to-NH White YPLL and mortality RR’s in the U.S. and in each of the 45 examined states and the District of Columbia with respect to cumulative COVID-19 deaths according to data from the National Center for Health Statistics as of December 30, 2020. States are ordered from top to bottom in descending order of the signed difference between the lower limit of the YPLL RR interval and the upper limit of the mortality RR interval. Values are displayed on the base 10 logarithmic scale. Interval endpoints above 20.0 and below 0.30 are truncated, with the actual values numerically annotated.

### 3.3. Results for Hispanics

Figure 4 displays a graphical comparison between the conservative 95% interval estimates of the percentage of total YPLL, intervals denoting the entire plausible range of the percentage of total deaths, and the percent population share for Hispanics in the U.S. and in each examined state. Nationally, COVID-19 deaths among Hispanics are approximately proportional to their percent population share: Hispanics comprise 18.5% of the U.S. population and 18.6%–19.1% of COVID-19 deaths. At the individual state level, Hispanic intervals of the percentage of total deaths are completely above the Hispanic percent population share in 11 of the examined states and completely below in 29 of the examined states. Hispanic intervals of the percentage of total deaths are completely above the NH White intervals of the percentage of total deaths in 4 of the examined states (California, D.C., New Mexico, and Texas) and are completely above all of the corresponding intervals for the other racial/ethnic groups in 2 of those states (California and Texas). Although COVID-19 deaths among Hispanics nationally are about in line with their percent population share, Hispanic COVID-19 decedent ages in the U.S. overall are skewed disproportionately toward younger ages to such a degree that the U.S. national Hispanic conservative 95% interval estimate of the percentage of total YPLL ([30.3% – 31.0%]) is completely above the U.S. national Hispanic interval of the percentage of total deaths. This national trend of Hispanics dying from COVID-19 at earlier ages relative to non-Hispanics is also widely observed at the individual state level, with the Hispanic conservative 95% interval estimates of the percentage of total YPLL completely above the Hispanic intervals of the percentage of total deaths in 38 of the examined states and completely below in none of the examined states. For example, in Illinois, Hispanics represent 17.5% of the population and comprise a commensurate 17.3%–18.0% of total deaths, but the Hispanic conservative 95% interval estimate of the percentage of total YPLL is [35.5% – 39.5%]. Hispanic conservative 95% interval estimates of the percentage of total YPLL are completely above the Hispanic percent population share in 34 out of the 46 examined states, greater than the corresponding number for the intervals of the percentages of total deaths, and completely below in only 1 state (New Mexico). Furthermore, Hispanic conservative 95% interval estimates of the percentage of total YPLL are completely above the corresponding intervals for NH Whites in 10 of the examined states and completely above all of the corresponding intervals for the other racial/ethnic groups in 7 of those states, each of which is greater than the corresponding number for the intervals of the percentage of total deaths. The direction of the disparity in fact reverses when using YPLL rather than death counts to measure mortality in 20 of the 29 states where the Hispanic intervals of the percentage of total deaths are completely below the Hispanic percent population share. For example, in Connecticut, Hispanics represent 16.9% of the population and only 9.5%–10.7% of total deaths, but the Hispanic conservative 95% interval estimate of the percentage of total YPLL is [20.9% – 30.7%]. We also note that the results for Hispanics in New Mexico are anomalous compared to the rest of the examined states, with both the Hispanic interval of the percentage of total deaths and the Hispanic conservative 95% interval estimate of the percentage of total YPLL markedly below the Hispanic percent population share: Hispanics represent nearly 50% of the population in New Mexico but comprise only 34.9%–35.8% of total deaths, and the Hispanic conservative 95% interval estimate of the percentage of total YPLL is [29.8% – 36.7%].

**Figure 4.**
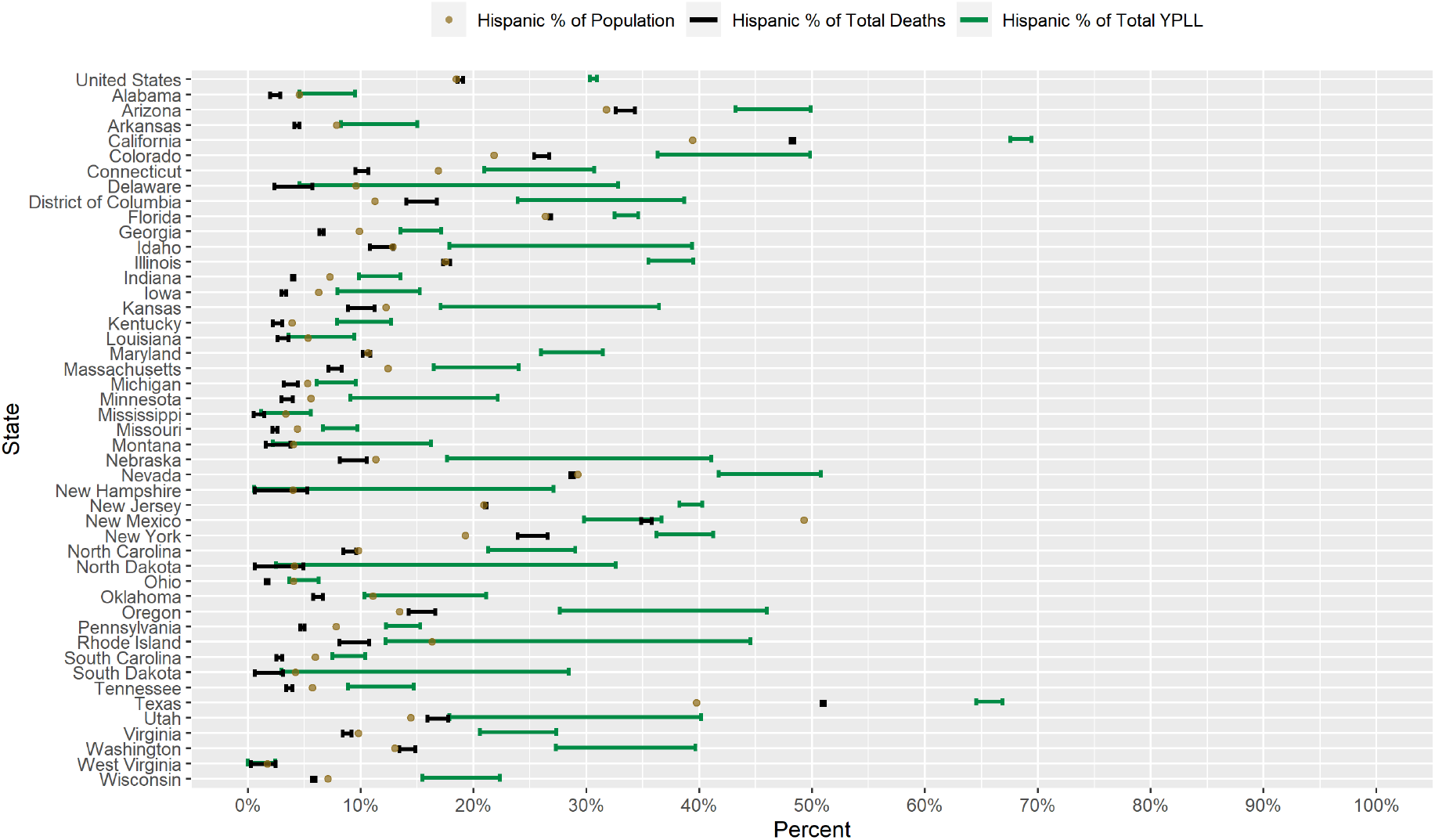
Conservative 95% interval estimates of the percentage of total YPLL before age 75, intervals denoting the entire plausible range for the percentage of total deaths, and the percent population shares for Hispanics in the U.S. and in each of the 45 examined states and the District of Columbia with respect to cumulative COVID-19 deaths according to data from the National Center for Health Statistics as of December 30, 2020.

Figure 5 presents a graphical comparison of the conservative 95% interval estimates of the age-adjusted Hispanic-to-NH White YPLL and mortality RR’s in the U.S. and in each examined state. The U.S. national conservative 95% interval estimate of the age-adjusted Hispanic-to-NH White mortality RR is [1.95 – 2.03], and conservative 95% interval estimates of the age-adjusted Hispanic-to-NH White mortality RR are completely above 1.0, 2.0, and 3.0 in 40, 21, and 6 of the examined states, respectively. However, Hispanic COVID-19 decedent ages in the U.S. overall are skewed disproportionately toward younger ages relative to NH Whites, after accounting for differences in the national population age distributions between Hispanics and NH Whites, to such a degree that the U.S. national conservative 95% interval estimate of the age-adjusted Hispanic-to-NH White YPLL RR is [2.90 – 3.02], completely above the U.S. national conservative 95% interval estimate of the age-adjusted Hispanic-to-NH White mortality RR. Indeed, the conservative 95% interval estimates of the age-adjusted Hispanic-to-NH White YPLL RR are completely above the conservative 95% interval estimates of the age-adjusted Hispanic-to-NH White mortality RR in 37 of the 46 examined states and are completely below in none of the examined states, reflecting a robust pattern across states of Hispanics dying from COVID-19 at earlier ages than NH Whites after accounting for differences in their respective state population age distributions. Moreover, conservative 95% interval estimates of the age-adjusted Hispanic-to-NH White YPLL RR are completely above 1.0, 2.0 and 3.0 in 42, 35, and 26 of the examined states, respectively, greater than the corresponding numbers for the age-adjusted Hispanic-to-NH White mortality RR’s. The conservative 95% interval estimates of the age-adjusted Hispanic-to-NH White YPLL RR are notably high and disconcerting in a sizable number of states: Illinois, Washington, and Wisconsin (completely above 5.0); California, North Carolina, and Oregon (completely above 6.0); Maryland (completely above 7.0); and D.C. (completely above 8.0).

**Figure 5.**
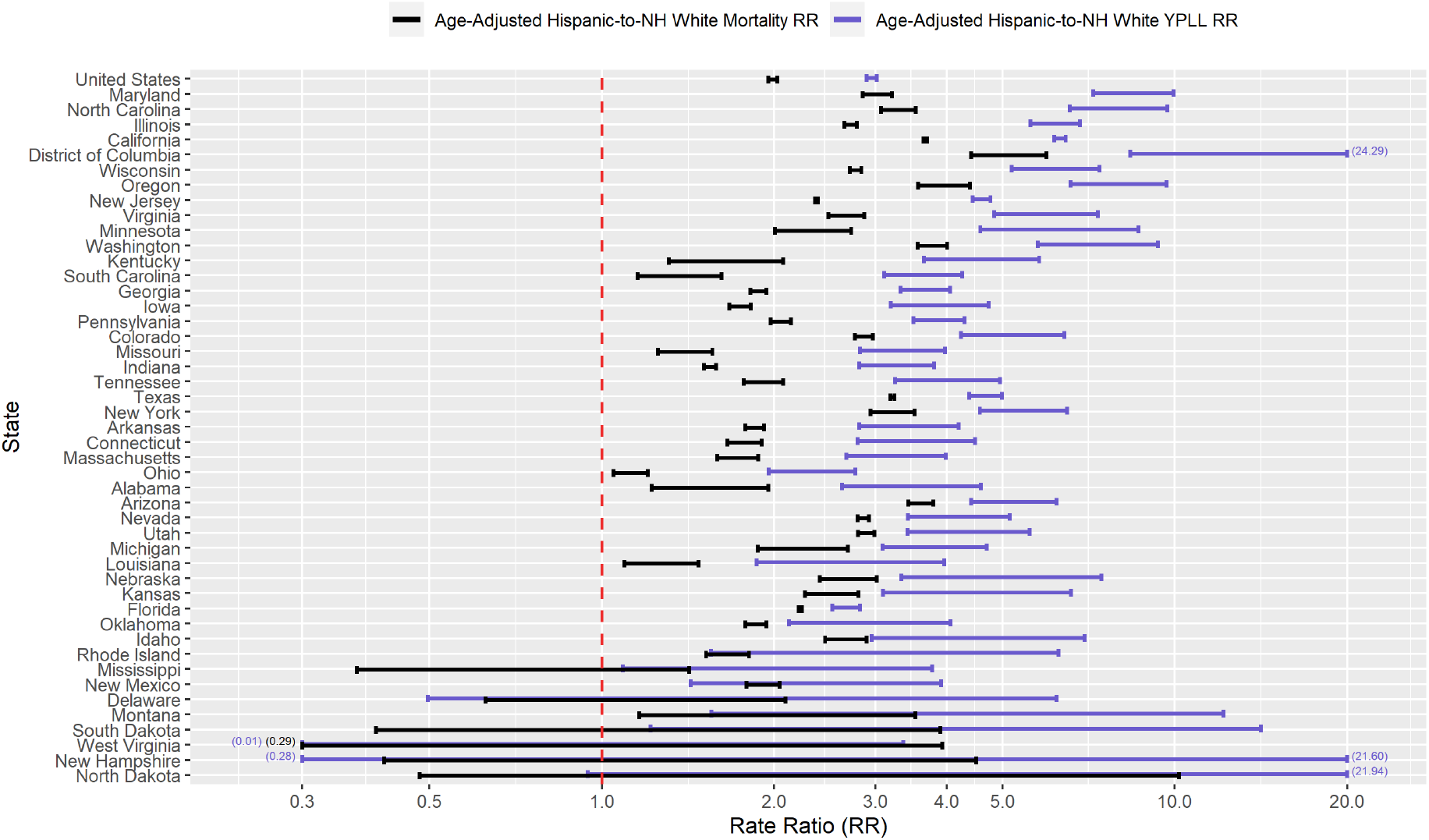
Conservative 95% interval estimates of the age-adjusted Hispanic-to-NH White YPLL and mortality RR’s in the U.S. and in each of the 45 examined states and the District of Columbia with respect to cumulative COVID-19 deaths according to data from the National Center for Health Statistics as of December 30, 2020. States are ordered from top to bottom in descending order of the signed difference between the lower limit of the YPLL RR interval and the upper limit of the mortality RR interval. Values are displayed on the base 10 logarithmic scale. Interval endpoints above 20.0 and below 0.30 are truncated, with the actual values numerically annotated.

### 3.4. Results for Non-Hispanic Asians

Figure 6 displays a graphical comparison between the conservative 95% interval estimates of the percentage of total YPLL, intervals denoting the entire plausible range of the percentage of total deaths, and the percent population share for NH Asians in the U.S. and in each examined state. Nationally, COVID-19 deaths among NH Asians are underrepresented, comprising only 3.6%–4.0% of COVID-19 deaths despite representing 6.3% of the U.S. population. Accordingly, NH Asian intervals of the percentage of total deaths are completely below the NH Asian percent population share in 32 of the examined states and completely above in only 1 examined state (Nevada). The U.S. national NH Asian conservative 95% interval estimate of the percentage of total YPLL ([3.8% – 4.4%]) is statistically indistinguishable from the U.S. national NH Asian interval of the percentage of total deaths. However, NH Asian conservative 95% interval estimates of the percentage of total YPLL contain the NH Asian percent population share in the vast majority of the examined states (*n* = 37), are completely above the NH Asian percent population share in 1 state (Minnesota), and are completely below the NH Asian percent population share in 8 states. Furthermore, in 3 of the examined states (Minnesota, Pennsylvania, and Wisconsin), the NH Asian conservative 95% interval estimates of the percentage of total YPLL are completely above the NH Asian intervals of the percentage of total deaths, indicating that NH Asians die from COVID-19 at statistically discernably earlier ages than individuals of other racial/ethnic groups in these states. Conversely, in 2 of the examined states (California and Nevada), the NH Asian conservative 95% interval estimates of the percentage of total YPLL are completely below the NH Asian intervals of the percentage of total deaths, indicating that NH Asians die from COVID-19 at statistically discernably older ages than individuals of other racial/ethnic groups in these states. In Minnesota, the direction of the disparity in fact reverses when using YPLL rather than death counts to measure mortality: NH Asians comprise 5.5% of the population and only 3.6%–4.5% of total deaths, but the conservative 95% interval estimate of the percentage of total YPLL is [8.0% – 15.1%]. The estimated ordered relationship between the NH Asian percent population share, percentage of total deaths, and percentage of total YPLL in California is unique among the 46 examined states: NH Asians represent 16.0% of the population but constitute only 11.6%–11.9% of total COVID-19 deaths. However, NH Asian decedent ages are skewed disproportionately toward older ages relative to the other racial/ethnic groups to such a degree that the conservative 95% interval estimate of the percentage of total YPLL is only [7.1% – 8.2%], completely below the NH Asian interval of the percentage of total deaths.

**Figure 6.**
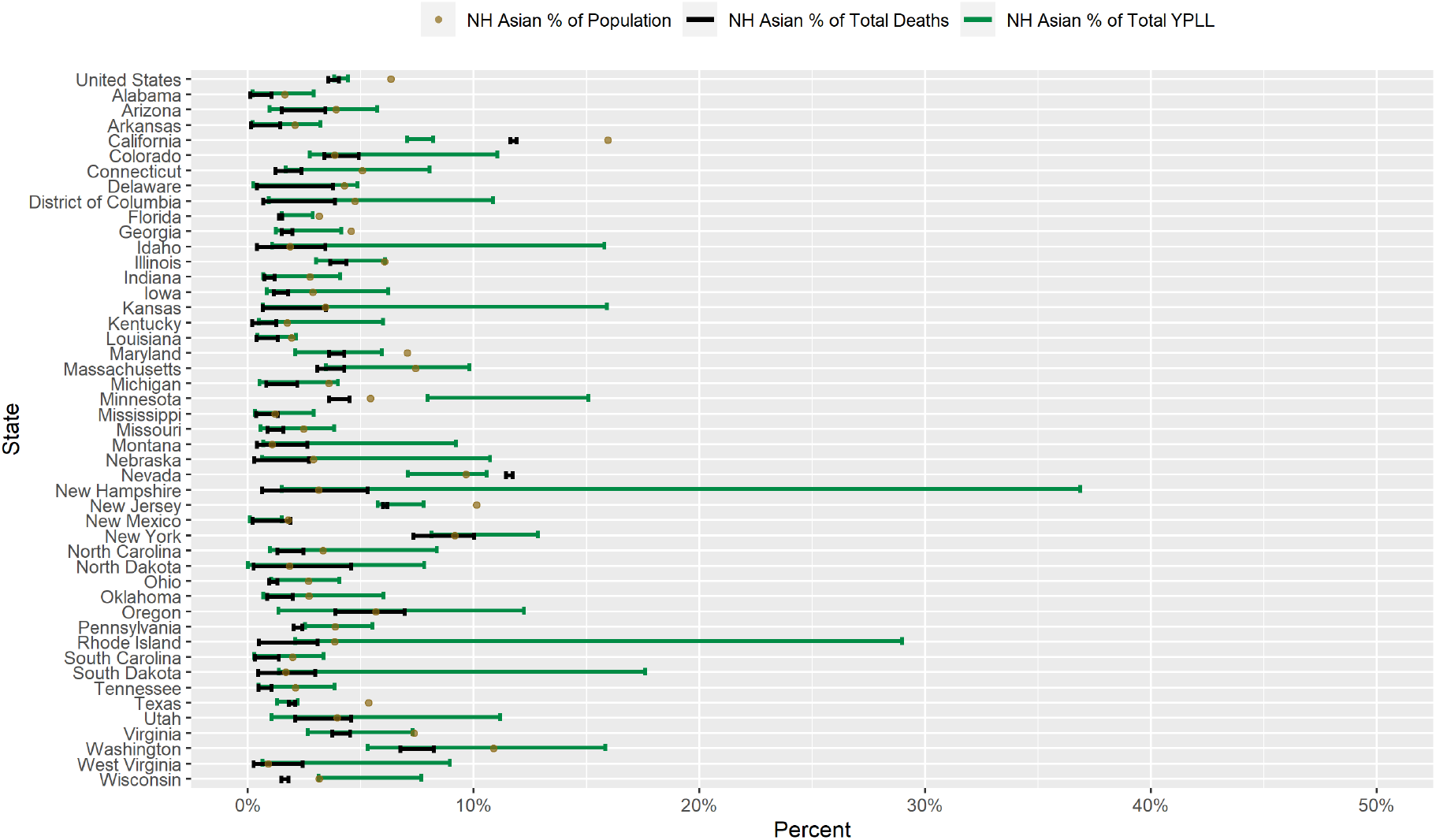
Conservative 95% interval estimates of the percentage of total YPLL before age 75, intervals denoting the entire plausible range for the percentage of total deaths, and the percent population shares for NH Asians in the U.S. and in each of the 45 examined states and the District of Columbia with respect to cumulative COVID-19 deaths according to data from the National Center for Health Statistics as of December 30, 2020.

Figure 7 presents a graphical comparison of the conservative 95% interval estimates of the age-adjusted NH Asian-to-NH White YPLL and mortality RR’s in the U.S. and in each examined state. The U.S. national conservative 95% interval estimate of the age-adjusted NH Asian-to-NH White mortality RR is [0.83 – 0.95], meaning that after accounting for differences in the national population age distributions between NH Asians and NH Whites, NH Asians actually die from COVID-19 at a rate that is statistically discernably lower than that of NH Whites. At the individual state level, however, the conservative 95% interval estimates of the age-adjusted NH Asian-to-NH White mortality RR are completely above 1.0 in 14 of the examined states and completely below 1.0 in only 4 of the examined states. Moreover, NH Asian decedent ages in the U.S. overall are skewed disproportionately toward younger ages relative to NH Whites, after accounting for differences in the national population age distributions between NH Asians and NH Whites, to such a degree that the U.S. national conservative 95% interval estimate of the age-adjusted NH Asian-to-NH White YPLL RR is [0.99 – 1.17], completely above the U.S. national conservative 95% interval estimate of the age-adjusted NH Asian-to-NH White mortality RR and also nearly representing a statistically discernable reversal in the direction of the age-adjusted NH Asian-NH White disparity in the COVID-19 mortality burden. At the individual state level, however, the conservative 95% interval estimates of the age-adjusted NH Asian-to-NH White YPLL RR are completely above the intervals of the age-adjusted NH Asian-to-NH White mortality RR in only 4 of the 46 examined states (California, Minnesota, New Jersey, and Wisconsin) and completely below in none of the examined states, suggesting that the age-adjusted national trend of NH Asians dying from COVID-19 at earlier ages than NH Whites is driven in large part by a small subset of states. Furthermore, conservative 95% interval estimates of the age-adjusted NH Asian-to-NH White YPLL RR are completely above 1.0 in 11 of the examined states, slightly below the corresponding number for the age-adjusted NH Asian-to-NH White mortality RR’s, but completely above 2.0 in 2 of the examined states (Minnesota and Wisconsin). In particular, the conservative 95% interval estimate of the age-adjusted NH Asian-to-NH White YPLL RR in Minnesota ([3.82 – 6.05]) stands out markedly among the examined states with the highest age-adjusted NH Asian-NH White disparity in COVID-19-attributable YPLL.

**Figure 7.**
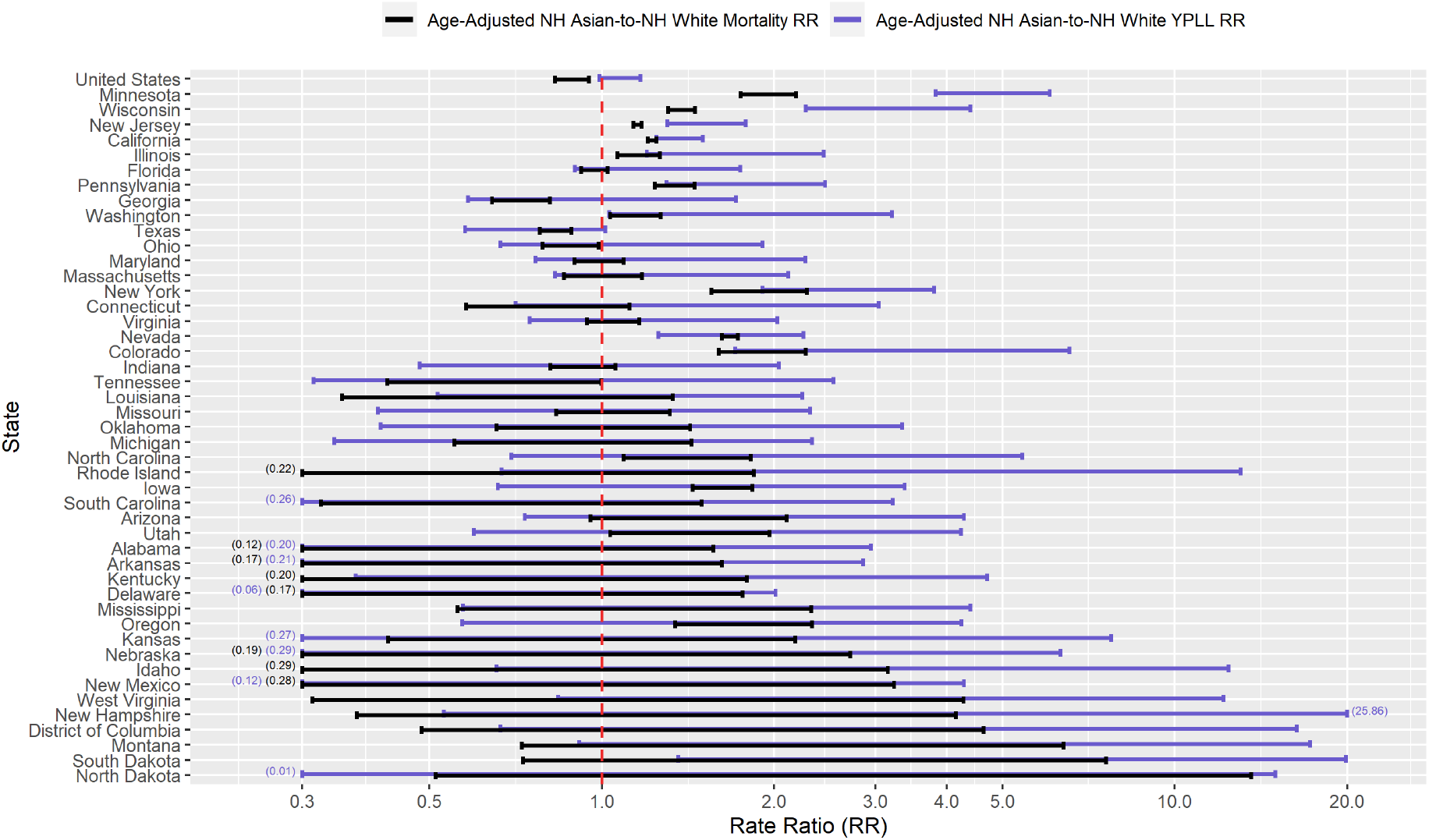
Conservative 95% interval estimates of the age-adjusted NH Asian-to-NH White YPLL and mortality RR’s in the U.S. and in each of the 45 examined states and the District of Columbia with respect to cumulative COVID-19 deaths according to data from the National Center for Health Statistics as of December 30, 2020. States are ordered from top to bottom in descending order of the signed difference between the lower limit of the YPLL RR interval and the upper limit of the mortality RR interval. Values are displayed on the base 10 logarithmic scale. Interval endpoints above 20.0 and below 0.30 are truncated, with the actual values numerically annotated.

### 3.5. Results for Non-Hispanic American Indian or Alaska Natives

Figure 8 displays a graphical comparison between the conservative 95% interval estimates of the percentage of total YPLL, intervals denoting the entire plausible range of the percentage of total deaths, and the percent population share for NH AIAN’s in the U.S. and in each examined state. Nationally, COVID-19 deaths among NH AIAN’s are overrepresented, comprising 1.1%–1.6% of COVID-19 deaths despite representing only 0.84% of the U.S. population. At the individual state level, NH AIAN intervals of the percentage of total deaths are completely above the NH AIAN population share in 12 of the examined states and completely below in only 1 state (Florida). However, NH AIAN COVID-19 decedent ages in the U.S. overall are skewed disproportionately toward younger ages to such a degree that the U.S. national NH AIAN conservative 95% interval estimate of the percentage of total YPLL is [2.0% – 2.6%], completely above the NH AIAN interval of the percentage of total deaths. Moreover, at the individual state level, the NH AIAN conservative 95% interval estimates of the percentage of total YPLL are completely above the NH AIAN intervals of the percentage of total deaths in 10 of the examined states and completely below in none of the examined states. For example, in South Dakota, NH AIAN’s represent 8.8% of the population and 10.6%–12.4% of total deaths, but the NH AIAN conservative 95% interval estimate of the percentage of total YPLL is an even greater [20.4% − 45.5%]. The NH AIAN conservative 95% interval estimates of the percentage of total YPLL are completely above the NH AIAN percent population share in 15 of the examined states, greater than the corresponding number for the NH AIAN intervals of the percentage of total deaths, and completely below in none of the examined states. The magnitudes of the disparities in the COVID-19 mortality burden for NH AIAN’s are shockingly immense in 3 of the examined states (Arizona, Montana, and New Mexico), with both the NH AIAN conservative 95% interval estimates of the percentage of total YPLL and the NH AIAN intervals of the percentage of total deaths exorbitantly exceeding the NH AIAN percent population share. For example, NH AIAN’s represent only 9.1% of the population of New Mexico but a stunning 34.8%–35.7% of total deaths, yet the interval estimate of the percentage of total YPLL is a staggering [48.5% – 57.0%].

**Figure 8.**
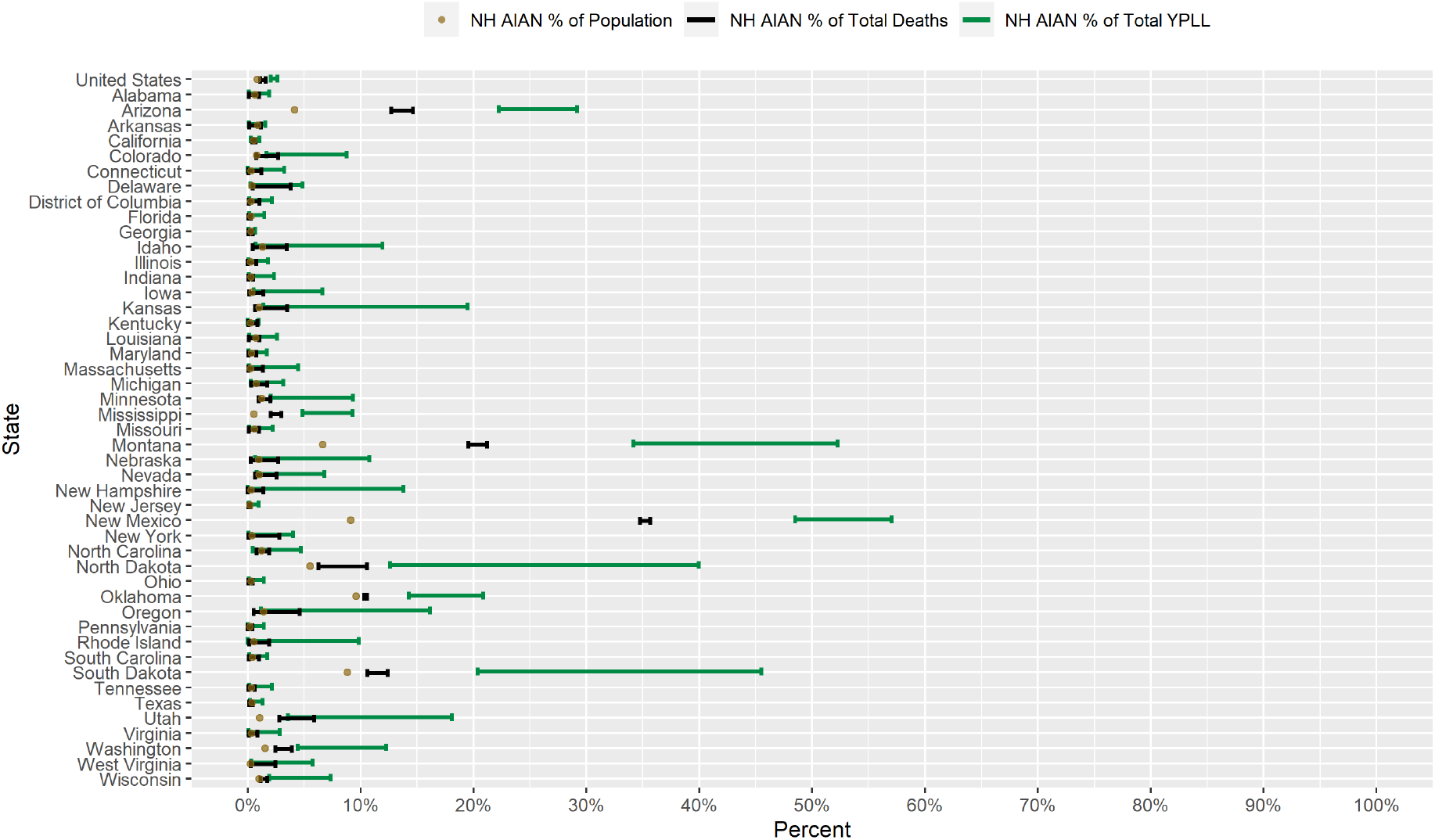
Conservative 95% interval estimates of the percentage of total YPLL before age 75, intervals denoting the entire plausible range for the percentage of total deaths, and the percent population shares for NH AIAN’s in the U.S. and in each of the 45 examined states and the District of Columbia with respect to cumulative COVID-19 deaths according to data from the National Center for Health Statistics as of December 30, 2020.

Figure 9 presents a graphical comparison of the conservative 95% interval estimates of the age-adjusted NH AIAN-to-NH White YPLL and mortality RR’s in the U.S. and in each examined state. The U.S. national conservative 95% interval estimate of the age-adjusted NH AIAN-to-NH White mortality RR is [1.82 – 2.70], and the conservative 95% interval estimates of the age-adjusted NH AIAN-to-NH White mortality RR are completely above 1.0, 2.0, and 3.0 in 14, 8, and 6 of the examined states, respectfully. However, NH AIAN COVID-19 decedent ages in the U.S. overall are skewed disproportionately toward younger ages relative to NH Whites, after accounting for differences in the national population age distributions between NH AIAN’s and NH Whites, to such a degree that the U.S. national conservative 95% interval estimate of the age-adjusted NH AIAN-to-NH White YPLL RR is [3.79 – 5.06], completely above the U.S. national conservative 95% interval estimate of the age-adjusted NH AIAN-to-NH White mortality RR. At the individual state level, the conservative 95% interval estimates of the age-adjusted NH AIAN-to-NH White YPLL RR are completely above the conservative 95% interval estimates of the age-adjusted NH Asian-to-NH White mortality RR in 7 of the 46 examined states and completely below in none of the examined states, suggesting that the age-adjusted national trend of NH AIAN’s dying from COVID-19 at younger ages relative to NH Whites is driven in large part by a small subset of states. Furthermore, conservative 95% interval estimates of the age-adjusted NH AIAN-to-NH White YPLL RR are completely above 1.0, 2.0, and 3.0 in 20, 14, and 11 of the examined states, respectfully, greater than the corresponding numbers for the age-adjusted NH AIAN-to-NH White mortality RR’s. The conservative 95% interval estimates of the age-adjusted NH AIAN-to-NH White YPLL RR in 4 of the examined states (which are among the 7 states where the conservative 95% interval estimates of the age-adjusted NH AIAN-to-NH White YPLL RR are completely above the conservative 95% interval estimates of the age-adjusted NH AIAN-to-NH White mortality RR) stand out starkly among the examined states with the highest age-adjusted NH AIAN-NH White disparities in COVID-19-attributable YPLL and are simply astronomical: Arizona ([14.79 – 23.70]), Mississippi ([17.97 – 34.13]), Montana ([11.14 – 22.46]), and New Mexico ([11.83 – 31.63]).

**Figure 9.**
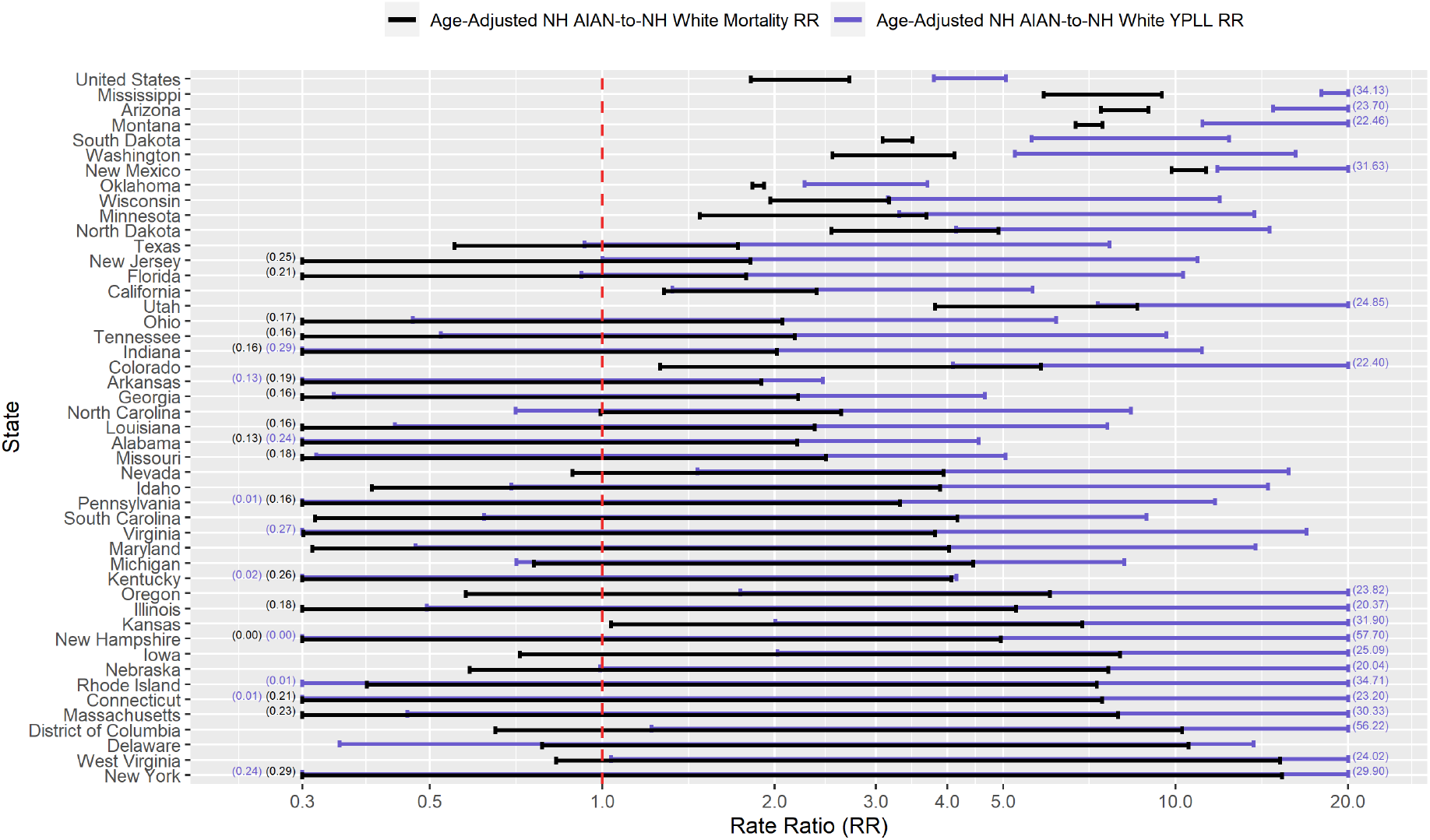
Conservative 95% interval estimates of the age-adjusted NH AIAN-to-NH White YPLL and mortality RR’s in the U.S. and in each of the 45 examined states and the District of Columbia with respect to cumulative COVID-19 deaths according to data from the National Center for Health Statistics as of December 30, 2020. States are ordered from top to bottom in descending order of the signed difference between the lower limit of the YPLL RR interval and the upper limit of the mortality RR interval. Values are displayed on the base 10 logarithmic scale. Interval endpoints above 20.0 and below 0.30 are truncated, with the actual values numerically annotated.

## 4. Discussion

Over 926,058 years of potential life before the age of 75 have been lost in the U.S. as a result of COVID-19, according to (lagged) NCHS data as of December 30, 2020, corresponding to 301,679 deaths. Proportionally scaling up this lower bound to the true number of COVID-19 deaths as of December 30, 2020 (342,577), as documented by the New York Times, results in an estimated lower bound of total U.S. COVID-19-attributable YPLL in excess of 1 million. However, the relative COVID-19 mortality burden by race/ethnicity has been far from equitable. Building upon the national comparative YPLL analysis by race/ethnicity by Bassett et al., we performed a corresponding state-by-state analysis, quantifying racial/ethnic disparities in COVID-19-attributable YPLL and death counts in 45 states and D.C. Because our YPLL analyses were performed at a more geographically precise level, they allowed for the identification of individual states where racial/ethnic disparities are particularly exacerbated and where younger non-NH White populations have been disproportionately devastated, as well as states that are exceptions to overarching national trends, which may aid in ascertaining important state-level characteristics that may be associated with the observed racial/ethnic disparities in COVID-19 mortality. Although a comprehensive investigation into the specific reasons as to why certain states possess higher degrees of racial/ethnic disparities in COVID-19-attributable mortality than others is outside the scope of this paper, our analyses serve as an initial exploratory tool to investigate why communities of color are harder hit from the U.S. COVID-19 epidemic in some states than in others.

Researchers have proposed multiple narratives to explain the vast disparities in the COVID-19 mortality burden by race/ethnicity. For example, Blacks and Hispanics in the U.S. have higher rates of underlying medical conditions such as diabetes, hypertension, and obesity [66,67], which have been established as definitive or probable risk factors of severe illness as a result of COVID-19 infection [68], and it has been hypothesized that the high prevalence of these conditions among Blacks and Hispanics in the U.S. contributes in large part to the observed disparities. Genetic factors have also been speculated as playing a role in the excess mortality experienced by Blacks in particular [69–71]. However, given that racial/ethnic disparities in the COVID-19 mortality burden reflect disparities in both the risk of COVID-19 infection and the risk of severe illness given COVID-19 infection, the underlying causes of the racial/ethnic disparities in the COVID-19 mortality burden are likely multi-faceted, comprising a complex and interactive combination of factors, including the prevalence of pre-existing comorbid conditions and potentially genetic factors. However, the wide geographical variation of the magnitudes of the estimated racial/ethnic disparities in the COVID-19 mortality burden across states suggests that they are driven in large part by social determinants of health [72] whose degree of association with race/ethnicity varies state to state, a perspective widely propounded in the scientific community [73–78]. In the context of COVID-19, social determinants of health that shape different experiences of individuals in different racial/ethnic groups may include differential access to healthcare, housing conditions, environmental conditions, and domain of employment [79]. For example, Blacks and Hispanics are overrepresented in many frontline essential industries [80–82], resulting in higher rates of COVID-19 exposure and potential subsequent mortality. Blacks and Hispanics also generally experience higher rates of industrial air pollution [83], which may increase the risk of COVID-19 mortality [84]. While an important limitation of our analyses is that they are descriptive in nature, the sheer magnitude of many of the estimated state-level disparities in our analyses speaks to the degree of urgency for communities of color in relation to the U.S. COVID-19 epidemic.

At the time of writing, two COVID-19 vaccines, one manufactured by Pfizer, Inc./BioNTech [85] and the other by ModernaTX, Inc. [86], have been granted emergency use authorization by the U.S. Food and Drug Administration, with several vaccine candidates in Phase III clinical trials. Vaccine distribution strategy has been the subject of rigorous public policy debate, with varying opinions on which segments of the population to prioritize for vaccination [87,88]. In any case, it is critical that communities of color particularly devastated by the U.S. COVID-19 epidemic have direct and equitable access to COVID-19 vaccines. Nevertheless, while vaccination serves as the most potent tool to fight COVID-19, both vaccine-based and non-pharmaceutical interventions should be pursued to prevent the further devastation of communities of color from the U.S. COVID-19 epidemic and to confront future public health crises.

## 5. Conclusions

In summary, we performed an extensive secondary analysis of U.S. national COVID-19 death counts stratified by state, race/ethnicity, and age group from the National Center for Health Statistics as of December 30, 2020, quantifying racial/ethnic disparities in COVID-19-attributable YPLL before age 75 in each of 45 states and D.C. Specifically, we quantified these disparities in YPLL through the estimation of percentages of total YPLL by race/ethnicity, contrasting them with their respective percent population shares, as well as age-adjusted YPLL RR’s for NH Blacks, Hispanics, NH Asians, and NH AIAN’s relative to NH Whites. Substantially complicating our analyses, however, were three sources of uncertainty in the data, namely, the administrative interval censoring of ages at death (precluding the exact calculation of YPLL), suppression of death counts between 1 and 9 within age intervals denoting age at death, and unknown race/ethnicity for a subset of COVID-19 deaths. We overcame these challenges in estimation by developing a novel adaptation of the MC simulation procedure proposed by Xu et al. that targets estimation of the extrema of the values of the estimands of interest that could theoretically be attained. However, a consequence of this conservative estimation strategy was wide interval estimates in scenarios for states and racial/ethnic groups corresponding to a high ratio of potential suppressed death counts to non-suppressed death counts and/or a comparatively large number of deaths with unknown race/ethnicity, which can reduce the power to detect racial/ethnic disparities when they exist and are small. Despite these challenges in estimation, our analyses revealed substantial racial/ethnic disparities in COVID-19-attributable YPLL before age 75 across U.S. states, with a prevailing pattern of NH Blacks and Hispanics experiencing disproportionately high COVID-19-attributable YPLL and NH Whites experiencing disproportionately low COVID-19-attributable YPLL. For comparison, we also calculated bounds for the corresponding percentages of total deaths and estimated the corresponding age-adjusted mortality RR’s in the 45 examined states and D.C., which revealed that racial/ethnic disparities in the COVID-19 mortality burden are generally greater in magnitude when measuring mortality in terms of YPLL compared to (age-irrespective) death counts, reflecting the greater intensity of the disparities at younger ages. The substantial state-to-state variation in the magnitudes of the estimated racial/ethnic disparities suggests that these disparities are driven in large part by social determinants of health encompassing domains such as healthcare access, housing conditions, environmental conditions, and domain of employment, whose degree of association with race/ethnicity varies by state. COVID-19 certainly didn’t cause racial/ethnic disparities in health outcomes, but it did highlight and bring unprecedented national attention to long-standing societal and health inequalities that many communities of color in the U.S. face. Only time will tell whether the U.S. COVID-19 epidemic catalyzes sustained efforts to reduce and eventually eliminate racial/ethnic disparities in health outcomes.

## Data Availability

Data referenced in the manuscript are available in the Supplemental Files.

## Supplementary Materials

File S1: Cumulative COVID-19 death counts in the U.S. stratified by state, race/ethnicity, and age group from the National Center for Health Statistics as of December 30, 2020, File S2: Cumulative COVID-19 death counts in the U.S. stratified by state, sex, and age group from the National Center for Health Statistics as of December 30, 2020, File S3: 2019 CDC WONDER estimates of the U.S. population stratified by state, race, ethnicity, and age group, Table S1: Conservative 95% interval estimates of total YPLL by race/ethnicity in the U.S. and in each examined state, Table S2: Intervals denoting the entire plausible range of total deaths by race/ethnicity in the U.S. and in each examined state, Table S3: Conservative 95% interval estimates of the percentage of total YPLL and intervals denoting the entire plausible range of the percentage of total deaths by race/ethnicity in the U.S. and in each examined state, Table S4: Conservative 95% interval estimates of age-adjusted YPLL and mortality rates by race/ethnicity in the U.S. and in each examined state, Table S5: Conservative 95% interval estimates of the age-adjusted YPLL and mortality RR’s for NH Blacks, Hispanics, NH Asians, and NH AIAN’s relative to NH Whites in the U.S. and in each examined state.

## Author Contributions

Conceptualization, J.J.X.; Data curation, J.J.X.; Formal analysis, J.J.X.; Methodology, J.J.X.; Software, J.J.X.; Visualization, J.J.X.; Writing – original draft, J.J.X.; Writing – review & editing, J.J.X., J.T.C., T.R.B., R.S.B., M.A.S. and C.M.R.

## Abbreviations

The following abbreviations are used in this manuscript:

CDC: Centers for Disease Control and Prevention
D.C.: District of Columbia
NCHS: National Center for Health Statistics
NH AIAN: Non-Hispanic American Indian or Alaska Native
NH Asian: Non-Hispanic Asian
NH Black: Non-Hispanic Black
NH White: Non-Hispanic White
RR: Rate Ratio
U.S.: United States
YPLL: Years of Potential Life Lost

