## Supplementary material for "Racial and Ethnic Disparities in Years of Potential Life Lost Attributable to COVID-19 in the United States: An Analysis of 45 States and the District of Columbia": Table S1

| State | NH White | NH Black | Hispanic | NH Asian | NH AIAN |
|---|---|---|---|---|---|
| US | [1,353,819 -- 1,377,390] | [775,987 -- 798,111] | [1,022,177 -- 1,044,441] | [128,910 -- 149,805] | [68,629 -- 89,077] |
| AL | [12,949 -- 14,528] | [14,595 -- 14,991] | [1,461 -- 1,580] | [69 -- 108] | [35 -- 71] |
| AZ | [10,192 -- 12,752] | [1,543 -- 2,205] | [22,176 -- 23,145] | [495 -- 1,122] | [11,318 -- 12,136] |
| AR | [7,488 -- 8,609] | [3,142 -- 3,316] | [1,307 -- 1,416] | [36 -- 54] | [12 -- 28] |
| CA | [21,213 -- 21,950] | [10,197 -- 10,756] | [99,053 -- 100,254] | [10,366 -- 10,915] | [366 -- 673] |
| CO | [5,770 -- 7,556] | [1,084 -- 1,208] | [6,731 -- 7,006] | [510 -- 613] | [305 -- 386] |
| CT | [6,831 -- 8,202] | [3,567 -- 3,823] | [3,427 -- 3,658] | [275 -- 380] | [0 -- 61] |
| DE | [1,554 -- 3,078] | [1,064 -- 1,173] | [203 -- 246] | [10 -- 20] | [10 -- 20] |
| DC | [281 -- 834] | [4,287 -- 4,493] | [1,937 -- 2,056] | [78 -- 101] | [10 -- 20] |
| FL | [29,642 -- 31,306] | [28,728 -- 29,256] | [30,640 -- 31,194] | [1,425 -- 1,554] | [132 -- 164] |
| GA | [17,140 -- 18,586] | [26,052 -- 26,563] | [7,388 -- 7,629] | [678 -- 774] | [30 -- 40] |
| ID | [2,392 -- 3,709] | [22 -- 45] | [904 -- 1,009] | [56 -- 81] | [35 -- 62] |
| IL | [20,147 -- 22,899] | [18,922 -- 19,553] | [25,097 -- 25,735] | [2,152 -- 2,505] | [36 -- 264] |
| IN | [17,084 -- 19,503] | [5,192 -- 5,477] | [2,759 -- 2,956] | [196 -- 272] | [22 -- 75] |
| IA | [9,235 -- 10,388] | [794 -- 876] | [1,185 -- 1,289] | [126 -- 174] | [79 -- 101] |
| KS | [4,969 -- 6,759] | [937 -- 1,116] | [1,983 -- 2,195] | [79 -- 185] | [164 -- 289] |
| KY | [8,828 -- 10,170] | [1,716 -- 1,872] | [1,077 -- 1,166] | [69 -- 97] | [0 -- 18] |
| LA | [10,948 -- 12,032] | [20,822 -- 21,299] | [1,329 -- 1,465] | [161 -- 231] | [55 -- 108] |
| MD | [6,218 -- 7,490] | [12,981 -- 13,365] | [7,827 -- 8,087] | [639 -- 760] | [23 -- 61] |
| MA | [13,739 -- 15,762] | [3,919 -- 4,439] | [4,211 -- 4,713] | [889 -- 1,310] | [22 -- 362] |
| MI | [21,204 -- 22,771] | [20,378 -- 21,232] | [2,958 -- 3,515] | [245 -- 718] | [132 -- 583] |
| MN | [7,429 -- 8,598] | [1,961 -- 2,102] | [1,367 -- 1,481] | [1,221 -- 1,337] | [311 -- 375] |
| MS | [8,499 -- 9,493] | [14,968 -- 15,369] | [324 -- 401] | [88 -- 158] | [1,346 -- 1,469] |
| MO | [14,636 -- 16,458] | [4,606 -- 4,843] | [1,619 -- 1,739] | [139 -- 198] | [35 -- 61] |
| MT | [1,852 -- 2,651] | [55 -- 75] | [105 -- 144] | [32 -- 48] | [1,577 -- 1,700] |
| NE | [4,101 -- 5,551] | [413 -- 494] | [1,464 -- 1,592] | [55 -- 91] | [55 -- 91] |
| NV | [4,195 -- 5,523] | [2,084 -- 2,235] | [7,731 -- 7,987] | [1,325 -- 1,466] | [150 -- 191] |
| NH | [802 -- 2,143] | [36 -- 70] | [12 -- 45] | [32 -- 65] | [0 -- 20] |
| NJ | [25,477 -- 26,484] | [20,227 -- 20,706] | [33,258 -- 33,791] | [5,013 -- 5,263] | [79 -- 101] |
| NM | [1,242 -- 2,532] | [69 -- 91] | [4,434 -- 4,644] | [12 -- 28] | [7,232 -- 7,464] |
| NY | [47,678 -- 59,498] | [55,669 -- 62,238] | [76,085 -- 82,646] | [17,130 -- 23,324] | [79 -- 5,966] |
| NC | [5,979 -- 7,035] | [6,641 -- 6,944] | [4,017 -- 4,244] | [190 -- 285] | [79 -- 167] |
| ND | [2,040 -- 3,475] | [22 -- 61] | [123 -- 170] | [0 -- 34] | [632 -- 729] |
| OH | [20,730 -- 21,718] | [7,029 -- 7,302] | [1,175 -- 1,279] | [329 -- 394] | [36 -- 55] |
| OK | [6,304 -- 8,532] | [897 -- 1,003] | [1,548 -- 1,669] | [104 -- 147] | [2,137 -- 2,292] |
| OR | [2,119 -- 2,609] | [113 -- 144] | [1,640 -- 1,761] | [81 -- 126] | [69 -- 98] |
| PA | [25,569 -- 27,175] | [12,144 -- 12,573] | [5,873 -- 6,151] | [1,215 -- 1,413] | [0 -- 81] |
| RI | [2,134 -- 3,260] | [465 -- 569] | [615 -- 740] | [113 -- 174] | [0 -- 48] |
| SC | [8,223 -- 9,374] | [11,263 -- 11,613] | [1,835 -- 1,943] | [69 -- 92] | [32 -- 48] |
| SD | [2,403 -- 3,233] | [56 -- 75] | [166 -- 194] | [80 -- 101] | [1,150 -- 1,259] |
| TN | [17,079 -- 19,214] | [9,377 -- 9,691] | [2,869 -- 3,029] | [157 -- 226] | [43 -- 82] |
| TX | [38,200 -- 42,254] | [22,343 -- 22,944] | [125,304 -- 126,456] | [2,512 -- 2,816] | [350 -- 547] |
| UT | [3,459 -- 4,465] | [36 -- 55] | [1,708 -- 1,835] | [105 -- 145] | [347 -- 411] |
| VA | [7,303 -- 8,791] | [7,364 -- 7,683] | [4,482 -- 4,710] | [584 -- 721] | [12 -- 77] |
| WA | [4,856 -- 6,385] | [472 -- 569] | [3,732 -- 3,929] | [733 -- 857] | [620 -- 716] |
| WV | [2,257 -- 3,078] | [88 -- 131] | [0 -- 10] | [20 -- 30] | [10 -- 20] |
| WI | [10,177 -- 11,933] | [2,016 -- 2,162] | [2,890 -- 3,055] | [593 -- 667] | [355 -- 421] |

**Table S1**: Conservative 95% interval estimates of total COVID-19-attributable YPLL before the age of 75 years for NH Whites, NH Blacks, Hispanics, NH Asians, and NH AIAN's in the U.S. and each examined state with respect to cumulative COVID-19 deaths according to data from the National Center for Health Statistics as of December 30, 2020.
