## Supplementary material for "Racial and Ethnic Disparities in Years of Potential Life Lost Attributable to COVID-19 in the United States: An Analysis of 45 States and the District of Columbia": Table S2

| State | NH White | NH Black | Hispanic | NH Asian | NH AIAN |
|---|---|---|---|---|---|
| US | [177,636 -- 179,096] | [51,054 -- 52,514] | [56,097 -- 57,557] | [10,725 -- 12,185] | [3,318 -- 4,778] |
| AL | [3,427 -- 3,470] | [1,654 -- 1,689] | [103 -- 152] | [6 -- 55] | [4 -- 53] |
| AZ | [3,283 -- 3,410] | [209 -- 344] | [2,270 -- 2,389] | [105 -- 240] | [884 -- 1,019] |
| AR | [2,440 -- 2,456] | [441 -- 449] | [129 -- 145] | [5 -- 45] | [4 -- 36] |
| CA | [6,865 -- 6,909] | [1,471 -- 1,531] | [10,418 -- 10,478] | [2,519 -- 2,579] | [81 -- 149] |
| CO | [2,472 -- 2,532] | [191 -- 243] | [989 -- 1,041] | [132 -- 192] | [29 -- 105] |
| CT | [3,738 -- 3,797] | [731 -- 790] | [483 -- 542] | [62 -- 121] | [2 -- 61] |
| DE | [499 -- 523] | [167 -- 191] | [17 -- 41] | [3 -- 27] | [3 -- 27] |
| DC | [105 -- 121] | [610 -- 626] | [123 -- 147] | [6 -- 34] | [1 -- 9] |
| FL | [10,182 -- 10,207] | [3,641 -- 3,658] | [5,188 -- 5,213] | [265 -- 298] | [7 -- 54] |
| GA | [4,561 -- 4,586] | [3,239 -- 3,264] | [541 -- 574] | [128 -- 168] | [3 -- 36] |
| ID | [1,012 -- 1,045] | [3 -- 36] | [129 -- 154] | [5 -- 41] | [5 -- 41] |
| IL | [8,327 -- 8,426] | [2,585 -- 2,668] | [2,415 -- 2,506] | [510 -- 609] | [4 -- 103] |
| IN | [6,154 -- 6,187] | [850 -- 875] | [289 -- 306] | [54 -- 87] | [3 -- 36] |
| IA | [3,588 -- 3,604] | [101 -- 125] | [117 -- 133] | [46 -- 70] | [6 -- 54] |
| KS | [2,062 -- 2,123] | [163 -- 232] | [228 -- 289] | [17 -- 89] | [17 -- 89] |
| KY | [2,635 -- 2,660] | [317 -- 342] | [69 -- 94] | [6 -- 39] | [2 -- 27] |
| LA | [3,179 -- 3,223] | [2,612 -- 2,670] | [159 -- 217] | [23 -- 81] | [5 -- 63] |
| MD | [2,688 -- 2,726] | [2,162 -- 2,200] | [579 -- 617] | [205 -- 243] | [4 -- 42] |
| MA | [7,421 -- 7,533] | [757 -- 869] | [660 -- 772] | [285 -- 397] | [4 -- 123] |
| MI | [6,725 -- 6,842] | [2,808 -- 2,933] | [323 -- 448] | [82 -- 223] | [32 -- 173] |
| MN | [4,113 -- 4,156] | [255 -- 298] | [142 -- 192] | [173 -- 216] | [45 -- 95] |
| MS | [2,281 -- 2,307] | [1,817 -- 1,851] | [22 -- 63] | [16 -- 57] | [86 -- 127] |
| MO | [5,142 -- 5,184] | [794 -- 828] | [137 -- 163] | [55 -- 97] | [5 -- 63] |
| MT | [716 -- 732] | [4 -- 25] | [15 -- 36] | [4 -- 25] | [185 -- 201] |
| NE | [1,533 -- 1,575] | [66 -- 110] | [147 -- 191] | [5 -- 49] | [5 -- 49] |
| NV | [1,173 -- 1,197] | [272 -- 288] | [725 -- 733] | [290 -- 298] | [16 -- 64] |
| NH | [598 -- 628] | [5 -- 35] | [4 -- 34] | [4 -- 34] | [0 -- 9] |
| NJ | [9,060 -- 9,077] | [2,843 -- 2,860] | [3,456 -- 3,481] | [985 -- 1,016] | [6 -- 37] |
| NM | [513 -- 544] | [6 -- 37] | [642 -- 659] | [4 -- 35] | [640 -- 657] |
| NY | [15,459 -- 16,449] | [8,147 -- 9,113] | [8,581 -- 9,531] | [2,631 -- 3,597] | [20 -- 1,010] |
| NC | [1,964 -- 1,999] | [1,045 -- 1,072] | [289 -- 328] | [45 -- 84] | [26 -- 65] |
| ND | [981 -- 1,029] | [3 -- 51] | [7 -- 55] | [3 -- 51] | [70 -- 118] |
| OH | [7,526 -- 7,543] | [1,242 -- 1,275] | [144 -- 169] | [86 -- 119] | [4 -- 38] |
| OK | [2,035 -- 2,067] | [162 -- 178] | [162 -- 186] | [24 -- 56] | [287 -- 295] |
| OR | [841 -- 867] | [7 -- 52] | [158 -- 184] | [43 -- 77] | [6 -- 51] |
| PA | [11,049 -- 11,102] | [2,127 -- 2,180] | [658 -- 711] | [288 -- 341] | [3 -- 56] |
| RI | [1,172 -- 1,197] | [81 -- 114] | [115 -- 152] | [7 -- 44] | [2 -- 27] |
| SC | [2,749 -- 2,774] | [1,675 -- 1,700] | [117 -- 142] | [15 -- 64] | [4 -- 45] |
| SD | [1,140 -- 1,156] | [4 -- 36] | [8 -- 42] | [6 -- 40] | [142 -- 166] |
| TN | [4,505 -- 4,539] | [1,138 -- 1,164] | [200 -- 234] | [28 -- 62] | [3 -- 37] |
| TX | [9,682 -- 9,760] | [2,699 -- 2,777] | [13,441 -- 13,519] | [486 -- 557] | [41 -- 119] |
| UT | [901 -- 917] | [5 -- 45] | [205 -- 229] | [27 -- 59] | [36 -- 76] |
| VA | [3,001 -- 3,040] | [1,236 -- 1,275] | [410 -- 449] | [182 -- 221] | [4 -- 43] |
| WA | [2,120 -- 2,165] | [89 -- 134] | [404 -- 447] | [203 -- 248] | [74 -- 117] |
| WV | [738 -- 755] | [18 -- 35] | [2 -- 19] | [2 -- 19] | [2 -- 19] |
| WI | [4,297 -- 4,321] | [342 -- 374] | [290 -- 306] | [76 -- 92] | [57 -- 89] |

**Table S2**: Intervals denoting the entire plausible range of total COVID-19 deaths for NH Whites, NH Blacks, Hispanics, NH Asians, and NH AIAN's in the U.S. and each examined state with respect to cumulative COVID-19 deaths according to data from the National Center for Health Statistics as of December 30, 2020.
