## Supplementary material for "Racial and Ethnic Disparities in Years of Potential Life Lost Attributable to COVID-19 in the United States: An Analysis of 45 States and the District of Columbia": Table S3

| | NH White | | NH Black | | Hispanic | | NH Asian | | NH AIAN | |
|---|---|---|---|---|---|---|---|---|---|---|
| State | % YPLL | % Deaths | % YPLL | % Deaths | % YPLL | % Deaths | % YPLL | % Deaths | % YPLL | % Deaths |
| US | [40.1 -- 40.8] | [58.9 -- 59.4] | [23.0 -- 23.6] | [16.9 -- 17.4] | [30.3 -- 31.0] | [18.6 -- 19.1] | [3.8 -- 4.4] | [3.6 -- 4.0] | [2.0 -- 2.6] | [1.1 -- 1.6] |
| AL | [41.0 -- 47.0] | [65.3 -- 66.1] | [45.6 -- 51.3] | [31.5 -- 32.2] | [4.6 -- 9.5] | [2.0 -- 2.9] | [0.21 -- 2.9] | [0.11 -- 1.0] | [0.11 -- 1.9] | [0.08 -- 1.0] |
| AZ | [19.9 -- 25.9] | [47.2 -- 49.0] | [3.0 -- 8.4] | [3.0 -- 4.9] | [43.2 -- 49.9] | [32.6 -- 34.3] | [0.97 -- 5.7] | [1.5 -- 3.4] | [22.2 -- 29.2] | [12.7 -- 14.6] |
| AR | [48.2 -- 61.6] | [77.7 -- 78.2] | [19.7 -- 27.3] | [14.0 -- 14.3] | [8.3 -- 15.0] | [4.1 -- 4.6] | [0.22 -- 3.2] | [0.16 -- 1.4] | [0.08 -- 1.5] | [0.13 -- 1.1] |
| CA | [14.4 -- 15.1] | [31.7 -- 31.9] | [6.9 -- 8.1] | [6.8 -- 7.1] | [67.5 -- 69.5] | [48.1 -- 48.4] | [7.1 -- 8.2] | [11.6 -- 11.9] | [0.25 -- 1.0] | [0.37 -- 0.69] |
| CO | [31.6 -- 45.6] | [63.4 -- 64.9] | [5.8 -- 13.2] | [4.9 -- 6.2] | [36.3 -- 49.8] | [25.4 -- 26.7] | [2.8 -- 11.1] | [3.4 -- 4.9] | [1.6 -- 8.8] | [0.74 -- 2.7] |
| CT | [41.4 -- 52.3] | [73.6 -- 74.8] | [22.0 -- 33.3] | [14.4 -- 15.6] | [20.9 -- 30.7] | [9.5 -- 10.7] | [1.7 -- 8.0] | [1.2 -- 2.4] | [0.00 -- 3.2] | [0.04 -- 1.2] |
| DE | [37.0 -- 69.4] | [69.8 -- 73.1] | [24.1 -- 54.9] | [23.4 -- 26.7] | [4.6 -- 32.8] | [2.4 -- 5.7] | [0.23 -- 4.8] | [0.42 -- 3.8] | [0.23 -- 4.8] | [0.42 -- 3.8] |
| DC | [3.4 -- 11.4] | [12.0 -- 13.8] | [52.3 -- 68.9] | [69.6 -- 71.4] | [23.9 -- 38.7] | [14.0 -- 16.8] | [0.94 -- 10.9] | [0.68 -- 3.9] | [0.12 -- 2.2] | [0.11 -- 1.0] |
| FL | [31.5 -- 33.6] | [52.5 -- 52.6] | [30.4 -- 32.2] | [18.8 -- 18.9] | [32.5 -- 34.6] | [26.8 -- 26.9] | [1.5 -- 2.9] | [1.4 -- 1.5] | [0.14 -- 1.5] | [0.04 -- 0.28] |
| GA | [31.4 -- 35.0] | [53.5 -- 53.8] | [47.8 -- 52.0] | [38.0 -- 38.3] | [13.5 -- 17.1] | [6.4 -- 6.7] | [1.2 -- 4.1] | [1.5 -- 2.0] | [0.06 -- 0.64] | [0.04 -- 0.42] |
| ID | [49.5 -- 77.1] | [84.8 -- 87.5] | [0.43 -- 8.8] | [0.25 -- 3.0] | [17.9 -- 39.4] | [10.8 -- 12.9] | [1.1 -- 15.8] | [0.42 -- 3.4] | [0.67 -- 11.9] | [0.42 -- 3.4] |
| IL | [28.6 -- 32.8] | [59.6 -- 60.4] | [26.8 -- 30.1] | [18.5 -- 19.1] | [35.5 -- 39.5] | [17.3 -- 18.0] | [3.0 -- 6.1] | [3.7 -- 4.4] | [0.05 -- 1.8] | [0.03 -- 0.74] |
| IN | [62.5 -- 69.7] | [83.2 -- 83.7] | [18.6 -- 24.3] | [11.5 -- 11.8] | [9.8 -- 13.5] | [3.9 -- 4.1] | [0.69 -- 4.1] | [0.73 -- 1.2] | [0.08 -- 2.3] | [0.04 -- 0.49] |
| IA | [62.7 -- 80.1] | [91.5 -- 91.9] | [5.3 -- 15.0] | [2.6 -- 3.2] | [7.9 -- 15.2] | [3.0 -- 3.4] | [0.84 -- 6.2] | [1.2 -- 1.8] | [0.53 -- 6.6] | [0.15 -- 1.4] |
| KS | [43.1 -- 66.9] | [80.4 -- 82.7] | [8.1 -- 27.7] | [6.4 -- 9.0] | [17.1 -- 36.4] | [8.9 -- 11.3] | [0.67 -- 15.9] | [0.66 -- 3.5] | [1.4 -- 19.5] | [0.66 -- 3.5] |
| KY | [67.0 -- 77.4] | [86.0 -- 86.8] | [12.6 -- 20.1] | [10.3 -- 11.2] | [7.9 -- 12.7] | [2.3 -- 3.1] | [0.50 -- 6.0] | [0.20 -- 1.3] | [0.00 -- 0.95] | [0.07 -- 0.88] |
| LA | [29.7 -- 34.7] | [52.6 -- 53.4] | [57.4 -- 64.6] | [43.2 -- 44.2] | [3.6 -- 9.4] | [2.6 -- 3.6] | [0.43 -- 2.1] | [0.38 -- 1.3] | [0.15 -- 2.6] | [0.08 -- 1.0] |
| MD | [20.5 -- 25.5] | [47.3 -- 48.0] | [43.2 -- 49.3] | [38.1 -- 38.7] | [26.0 -- 31.5] | [10.2 -- 10.9] | [2.1 -- 5.9] | [3.6 -- 4.3] | [0.07 -- 1.7] | [0.07 -- 0.74] |
| MA | [54.0 -- 62.7] | [80.1 -- 81.3] | [15.4 -- 22.9] | [8.2 -- 9.4] | [16.5 -- 24.0] | [7.1 -- 8.3] | [3.5 -- 9.8] | [3.1 -- 4.3] | [0.09 -- 4.5] | [0.04 -- 1.3] |
| MI | [43.9 -- 48.6] | [66.4 -- 67.6] | [42.4 -- 47.4] | [27.7 -- 29.0] | [6.1 -- 9.6] | [3.2 -- 4.4] | [0.51 -- 4.0] | [0.81 -- 2.2] | [0.27 -- 3.1] | [0.32 -- 1.7] |
| MN | [49.1 -- 62.8] | [86.0 -- 86.9] | [12.9 -- 21.7] | [5.3 -- 6.2] | [9.1 -- 22.2] | [3.0 -- 4.0] | [8.0 -- 15.1] | [3.6 -- 4.5] | [2.0 -- 9.3] | [0.94 -- 2.0] |
| MS | [30.7 -- 35.9] | [53.4 -- 54.1] | [54.4 -- 61.2] | [42.6 -- 43.4] | [1.2 -- 5.6] | [0.52 -- 1.5] | [0.32 -- 2.9] | [0.37 -- 1.3] | [4.8 -- 9.3] | [2.0 -- 3.0] |
| MO | [61.7 -- 71.0] | [82.9 -- 83.6] | [19.1 -- 25.3] | [12.8 -- 13.4] | [6.6 -- 9.7] | [2.2 -- 2.6] | [0.57 -- 3.8] | [0.89 -- 1.6] | [0.14 -- 2.2] | [0.08 -- 1.0] |
| MT | [40.3 -- 58.6] | [75.5 -- 77.2] | [1.2 -- 13.8] | [0.42 -- 2.6] | [2.2 -- 16.2] | [1.6 -- 3.8] | [0.68 -- 9.2] | [0.42 -- 2.6] | [34.1 -- 52.3] | [19.5 -- 21.2] |
| NE | [48.3 -- 72.5] | [84.9 -- 87.3] | [4.8 -- 24.6] | [3.7 -- 6.1] | [17.6 -- 41.1] | [8.1 -- 10.6] | [0.63 -- 10.7] | [0.28 -- 2.7] | [0.64 -- 10.8] | [0.28 -- 2.7] |
| NV | [22.6 -- 31.8] | [46.2 -- 47.2] | [11.2 -- 17.1] | [10.7 -- 11.4] | [41.7 -- 50.8] | [28.6 -- 28.9] | [7.1 -- 10.6] | [11.4 -- 11.7] | [0.79 -- 6.8] | [0.63 -- 2.5] |
| NH | [52.5 -- 95.9] | [93.1 -- 97.8] | [1.6 -- 38.6] | [0.78 -- 5.5] | [0.57 -- 27.1] | [0.62 -- 5.3] | [1.5 -- 36.9] | [0.62 -- 5.3] | [0.00 -- 13.8] | [0.00 -- 1.4] |
| NJ | [29.3 -- 30.8] | [55.1 -- 55.2] | [23.2 -- 24.6] | [17.3 -- 17.4] | [38.3 -- 40.2] | [21.0 -- 21.2] | [5.8 -- 7.8] | [6.0 -- 6.2] | [0.09 -- 0.97] | [0.04 -- 0.23] |
| NM | [8.4 -- 17.4] | [27.9 -- 29.5] | [0.45 -- 5.3] | [0.33 -- 2.0] | [29.8 -- 36.7] | [34.9 -- 35.8] | [0.08 -- 1.5] | [0.22 -- 1.9] | [48.5 -- 57.0] | [34.8 -- 35.7] |
| NY | [22.8 -- 28.4] | [43.1 -- 45.9] | [26.5 -- 31.7] | [22.7 -- 25.4] | [36.2 -- 41.2] | [23.9 -- 26.6] | [8.1 -- 12.9] | [7.3 -- 10.0] | [0.04 -- 4.0] | [0.06 -- 2.8] |
| NC | [31.4 -- 38.6] | [57.5 -- 58.6] | [34.6 -- 40.9] | [30.6 -- 31.4] | [21.3 -- 29.0] | [8.5 -- 9.6] | [1.0 -- 8.4] | [1.3 -- 2.5] | [0.42 -- 4.7] | [0.76 -- 1.9] |
| ND | [43.2 -- 80.1] | [87.9 -- 92.2] | [0.44 -- 14.0] | [0.27 -- 4.6] | [2.5 -- 32.6] | [0.63 -- 4.9] | [0.01 -- 7.8] | [0.27 -- 4.6] | [12.6 -- 40.0] | [6.3 -- 10.6] |
| OH | [65.9 -- 71.1] | [83.2 -- 83.4] | [22.3 -- 27.6] | [13.7 -- 14.1] | [3.7 -- 6.3] | [1.6 -- 1.9] | [1.0 -- 4.0] | [0.95 -- 1.3] | [0.11 -- 1.4] | [0.04 -- 0.42] |
| OK | [43.9 -- 61.5] | [73.0 -- 74.1] | [6.0 -- 12.9] | [5.8 -- 6.4] | [10.4 -- 21.1] | [5.8 -- 6.7] | [0.70 -- 6.0] | [0.86 -- 2.0] | [14.3 -- 20.9] | [10.3 -- 10.6] |
| OR | [35.5 -- 56.1] | [75.9 -- 78.2] | [1.9 -- 22.4] | [0.63 -- 4.7] | [27.6 -- 46.0] | [14.3 -- 16.6] | [1.4 -- 12.2] | [3.9 -- 6.9] | [1.1 -- 16.1] | [0.54 -- 4.6] |
| PA | [53.6 -- 58.2] | [77.9 -- 78.3] | [25.4 -- 29.6] | [15.0 -- 15.4] | [12.2 -- 15.3] | [4.6 -- 5.0] | [2.5 -- 5.5] | [2.0 -- 2.4] | [0.00 -- 1.4] | [0.02 -- 0.39] |
| RI | [40.4 -- 71.4] | [82.7 -- 84.4] | [8.8 -- 36.2] | [5.7 -- 8.0] | [12.2 -- 44.6] | [8.1 -- 10.7] | [2.1 -- 29.0] | [0.49 -- 3.1] | [0.00 -- 9.8] | [0.14 -- 1.9] |
| SC | [33.9 -- 41.1] | [59.6 -- 60.1] | [46.8 -- 54.9] | [36.3 -- 36.8] | [7.5 -- 10.4] | [2.5 -- 3.1] | [0.28 -- 3.4] | [0.32 -- 1.4] | [0.13 -- 1.7] | [0.09 -- 0.97] |
| SD | [42.5 -- 67.9] | [85.3 -- 86.5] | [0.95 -- 13.4] | [0.30 -- 2.7] | [3.0 -- 28.5] | [0.60 -- 3.1] | [1.4 -- 17.6] | [0.45 -- 3.0] | [20.4 -- 45.5] | [10.6 -- 12.4] |
| TN | [53.4 -- 60.1] | [76.2 -- 76.7] | [28.9 -- 33.4] | [19.2 -- 19.7] | [8.9 -- 14.7] | [3.4 -- 4.0] | [0.49 -- 3.9] | [0.47 -- 1.0] | [0.13 -- 2.2] | [0.05 -- 0.63] |
| TX | [19.7 -- 21.8] | [36.6 -- 36.9] | [11.5 -- 13.0] | [10.2 -- 10.5] | [64.5 -- 66.8] | [50.8 -- 51.1] | [1.3 -- 2.2] | [1.8 -- 2.1] | [0.18 -- 1.3] | [0.16 -- 0.45] |
| UT | [36.1 -- 63.2] | [70.0 -- 71.2] | [0.35 -- 6.6] | [0.39 -- 3.5] | [17.9 -- 40.2] | [15.9 -- 17.8] | [1.1 -- 11.2] | [2.1 -- 4.6] | [3.5 -- 18.1] | [2.8 -- 5.9] |
| VA | [33.6 -- 40.9] | [61.5 -- 62.3] | [33.7 -- 40.2] | [25.3 -- 26.1] | [20.6 -- 27.4] | [8.4 -- 9.2] | [2.7 -- 7.3] | [3.7 -- 4.5] | [0.06 -- 2.8] | [0.08 -- 0.88] |
| WA | [35.3 -- 49.3] | [70.5 -- 72.0] | [3.4 -- 13.1] | [3.0 -- 4.5] | [27.3 -- 39.7] | [13.4 -- 14.9] | [5.3 -- 15.8] | [6.8 -- 8.3] | [4.4 -- 12.3] | [2.5 -- 3.9] |
| WV | [79.8 -- 95.8] | [94.7 -- 96.9] | [2.8 -- 16.4] | [2.3 -- 4.5] | [0.01 -- 2.4] | [0.26 -- 2.4] | [0.64 -- 9.0] | [0.26 -- 2.4] | [0.33 -- 5.8] | [0.26 -- 2.4] |
| WI | [55.3 -- 66.3] | [84.2 -- 84.7] | [10.9 -- 21.2] | [6.7 -- 7.3] | [15.5 -- 22.3] | [5.7 -- 6.0] | [3.1 -- 7.7] | [1.5 -- 1.8] | [1.9 -- 7.4] | [1.1 -- 1.7] |

**Table S3**: Conservative 95% interval estimates of the percentage of total COVID-19-attributable YPLL before the age of 75 years and intervals denoting the entire plausible range of the percentage of total COVID-19 deaths for NH Whites, NH Blacks, Hispanics, NH Asians, and NH AIAN's in the U.S. and each examined state with respect to cumulative COVID-19 deaths according to data from the National Center for Health Statistics as of December 30, 2020.
