## Supplementary material for "Racial and Ethnic Disparities in Years of Potential Life Lost Attributable to COVID-19 in the United States: An Analysis of 45 States and the District of Columbia": Table S4

| State | NH White YPLL | NH White Mortality | NH Black YPLL | NH Black Mortality | Hispanic YPLL | Hispanic Mortality | NH Asian YPLL | NH Asian Mortality | NH AIAN YPLL | NH AIAN Mortality |
|---|---|---|---|---|---|---|---|---|---|---|
| US | [63.3 -- 64.5] | [7.4 -- 7.4] | [182.7 -- 188.0] | [14.2 -- 14.7] | [186.7 -- 191.2] | [14.5 -- 15.0] | [63.8 -- 74.2] | [6.2 -- 7.0] | [244.0 -- 321.0] | [13.5 -- 19.9] |
| AL | [36.6 -- 41.8] | [8.9 -- 9.0] | [115.8 -- 122.7] | [15.7 -- 16.1] | [108.7 -- 169.3] | [11.0 -- 17.4] | [8.3 -- 109.4] | [1.1 -- 13.9] | [10.2 -- 169.2] | [1.2 -- 19.4] |
| AZ | [22.0 -- 28.9] | [5.4 -- 5.7] | [52.8 -- 122.0] | [10.0 -- 15.6] | [126.5 -- 137.6] | [19.6 -- 20.6] | [21.0 -- 95.4] | [5.5 -- 11.4] | [423.3 -- 524.4] | [42.4 -- 48.8] |
| AR | [31.8 -- 37.1] | [9.1 -- 9.2] | [73.7 -- 84.8] | [13.1 -- 13.4] | [103.0 -- 135.2] | [16.4 -- 17.5] | [7.7 -- 92.1] | [1.5 -- 14.8] | [4.7 -- 78.8] | [1.7 -- 17.3] |
| CA | [12.6 -- 13.0] | [3.4 -- 3.4] | [42.7 -- 49.1] | [7.4 -- 7.7] | [79.9 -- 81.4] | [12.3 -- 12.4] | [16.1 -- 19.0] | [4.1 -- 4.2] | [17.2 -- 71.9] | [4.3 -- 8.0] |
| CO | [13.4 -- 18.2] | [6.2 -- 6.4] | [50.5 -- 88.3] | [13.0 -- 16.3] | [76.3 -- 86.8] | [17.6 -- 18.5] | [30.5 -- 89.2] | [10.2 -- 14.1] | [73.3 -- 304.2] | [8.0 -- 36.2] |
| CT | [23.2 -- 29.3] | [10.5 -- 10.8] | [102.5 -- 149.0] | [26.6 -- 28.8] | [80.7 -- 105.5] | [17.9 -- 20.1] | [20.3 -- 72.0] | [6.3 -- 11.8] | [0.47 -- 555.6] | [2.3 -- 78.5] |
| DE | [22.4 -- 53.4] | [5.9 -- 6.4] | [49.1 -- 100.2] | [10.0 -- 11.1] | [27.8 -- 140.2] | [4.1 -- 12.3] | [3.0 -- 45.9] | [1.1 -- 10.4] | [18.8 -- 320.9] | [5.1 -- 61.7] |
| DC | [15.8 -- 36.6] | [5.5 -- 6.3] | [140.6 -- 171.5] | [19.7 -- 20.3] | [298.7 -- 394.1] | [27.5 -- 33.5] | [22.6 -- 262.2] | [3.0 -- 26.0] | [42.6 -- 963.9] | [4.1 -- 58.4] |
| FL | [21.9 -- 23.6] | [5.1 -- 5.2] | [91.6 -- 94.7] | [14.4 -- 14.5] | [59.2 -- 62.0] | [11.4 -- 11.5] | [20.9 -- 38.5] | [4.7 -- 5.3] | [21.5 -- 227.6] | [1.1 -- 9.1] |
| GA | [28.1 -- 30.8] | [7.4 -- 7.5] | [81.4 -- 85.7] | [14.4 -- 14.6] | [101.5 -- 114.8] | [13.6 -- 14.4] | [17.8 -- 48.8] | [4.8 -- 6.0] | [10.4 -- 132.4] | [1.2 -- 16.3] |
| ID | [15.4 -- 24.7] | [6.6 -- 6.9] | [21.3 -- 347.9] | [5.9 -- 83.1] | [71.2 -- 110.1] | [16.8 -- 19.2] | [15.7 -- 197.2] | [2.0 -- 20.9] | [16.9 -- 231.8] | [2.7 -- 25.7] |
| IL | [23.0 -- 26.6] | [8.4 -- 8.6] | [109.1 -- 119.0] | [17.2 -- 17.7] | [148.0 -- 158.3] | [22.7 -- 23.6] | [31.5 -- 56.5] | [9.1 -- 10.7] | [13.0 -- 472.3] | [1.5 -- 44.6] |
| IN | [30.0 -- 34.6] | [10.4 -- 10.5] | [92.7 -- 111.3] | [19.8 -- 20.3] | [96.7 -- 115.2] | [15.8 -- 16.5] | [16.5 -- 61.8] | [8.5 -- 11.0] | [10.4 -- 338.0] | [1.6 -- 21.0] |
| IA | [32.0 -- 36.5] | [10.5 -- 10.6] | [94.4 -- 167.3] | [21.0 -- 23.2] | [115.0 -- 153.4] | [17.7 -- 19.2] | [24.2 -- 109.6] | [15.3 -- 19.4] | [71.9 -- 817.1] | [7.7 -- 84.2] |
| KS | [21.1 -- 29.4] | [7.6 -- 7.9] | [61.5 -- 162.8] | [14.3 -- 19.1] | [89.3 -- 141.5] | [17.8 -- 21.3] | [7.8 -- 165.9] | [3.3 -- 16.5] | [56.9 -- 687.9] | [8.1 -- 52.2] |
| KY | [21.6 -- 25.2] | [6.5 -- 6.6] | [51.7 -- 73.6] | [12.2 -- 13.2] | [90.7 -- 126.8] | [8.6 -- 13.5] | [9.0 -- 103.1] | [1.3 -- 11.7] | [0.21 -- 91.1] | [1.7 -- 26.7] |
| LA | [37.3 -- 41.3] | [10.3 -- 10.4] | [150.5 -- 164.3] | [22.9 -- 23.4] | [76.1 -- 149.5] | [11.4 -- 15.1] | [20.9 -- 84.7] | [3.7 -- 13.6] | [17.7 -- 287.4] | [1.7 -- 24.1] |
| MD | [17.9 -- 22.2] | [6.8 -- 6.9] | [70.0 -- 78.3] | [14.9 -- 15.2] | [157.6 -- 179.2] | [19.8 -- 21.9] | [16.7 -- 41.2] | [6.2 -- 7.4] | [10.3 -- 251.6] | [2.2 -- 27.4] |
| MA | [23.9 -- 28.0] | [11.4 -- 11.6] | [83.7 -- 115.9] | [21.7 -- 24.8] | [73.8 -- 96.4] | [18.5 -- 21.4] | [22.9 -- 51.2] | [10.0 -- 13.4] | [12.9 -- 731.3] | [2.7 -- 90.7] |
| MI | [25.1 -- 27.1] | [7.4 -- 7.6] | [154.9 -- 168.6] | [24.7 -- 25.8] | [82.7 -- 119.0] | [14.1 -- 19.9] | [9.2 -- 59.2] | [4.2 -- 10.6] | [19.2 -- 206.7] | [5.7 -- 33.0] |
| MN | [14.8 -- 17.5] | [7.5 -- 7.6] | [76.6 -- 100.2] | [18.3 -- 21.6] | [79.1 -- 129.5] | [15.3 -- 20.5] | [66.2 -- 91.2] | [13.3 -- 16.4] | [56.8 -- 206.5] | [11.3 -- 27.6] |
| MS | [47.3 -- 53.2] | [11.3 -- 11.5] | [142.8 -- 155.8] | [22.0 -- 22.4] | [57.1 -- 180.6] | [4.3 -- 16.1] | [30.2 -- 210.1] | [6.4 -- 26.2] | [942.4 -- 1,636.5] | [67.8 -- 106.9] |
| MO | [27.5 -- 31.4] | [8.9 -- 9.0] | [71.6 -- 85.2] | [15.0 -- 15.6] | [87.6 -- 110.9] | [11.2 -- 13.9] | [12.7 -- 64.3] | [7.4 -- 11.7] | [9.8 -- 140.1] | [1.6 -- 21.9] |
| MT | [17.9 -- 27.0] | [6.4 -- 6.6] | [85.8 -- 998.4] | [7.5 -- 171.0] | [40.0 -- 222.0] | [7.6 -- 22.6] | [24.0 -- 320.6] | [4.8 -- 41.0] | [294.7 -- 411.5] | [44.1 -- 47.7] |
| NE | [25.6 -- 35.3] | [8.4 -- 8.7] | [52.9 -- 190.8] | [13.1 -- 19.7] | [116.1 -- 193.8] | [20.7 -- 25.3] | [10.1 -- 164.6] | [1.7 -- 22.8] | [34.4 -- 520.3] | [5.1 -- 64.0] |
| NV | [22.8 -- 33.0] | [6.0 -- 6.3] | [80.9 -- 103.9] | [12.8 -- 13.4] | [111.3 -- 118.8] | [17.5 -- 17.7] | [40.6 -- 52.5] | [10.1 -- 10.5] | [47.5 -- 367.2] | [5.5 -- 23.9] |
| NH | [5.5 -- 17.3] | [4.1 -- 4.4] | [19.4 -- 307.5] | [4.7 -- 45.4] | [5.1 -- 124.8] | [1.9 -- 18.6] | [9.1 -- 146.9] | [1.7 -- 17.1] | [0.00 -- 335.6] | [0.00 -- 20.4] |
| NJ | [45.1 -- 47.3] | [13.2 -- 13.2] | [173.8 -- 181.1] | [29.2 -- 29.4] | [208.6 -- 216.2] | [31.2 -- 31.5] | [61.0 -- 81.1] | [15.0 -- 15.5] | [46.1 -- 497.9] | [3.4 -- 23.9] |
| NM | [13.7 -- 34.9] | [4.1 -- 4.6] | [16.1 -- 171.3] | [1.6 -- 12.7] | [49.0 -- 54.7] | [8.3 -- 8.5] | [3.9 -- 61.4] | [1.3 -- 13.4] | [406.8 -- 439.5] | [45.4 -- 46.9] |
| NY | [39.9 -- 51.1] | [10.6 -- 11.4] | [195.4 -- 230.6] | [31.5 -- 35.3] | [232.2 -- 259.7] | [33.5 -- 37.4] | [96.9 -- 152.2] | [17.7 -- 24.2] | [12.1 -- 1,198.2] | [3.3 -- 163.0] |
| NC | [8.0 -- 9.5] | [2.5 -- 2.5] | [29.7 -- 32.6] | [5.8 -- 6.0] | [61.4 -- 77.9] | [7.7 -- 8.7] | [6.5 -- 43.6] | [2.8 -- 4.5] | [6.5 -- 67.9] | [2.5 -- 6.4] |
| ND | [31.5 -- 54.4] | [12.5 -- 13.4] | [15.9 -- 501.0] | [6.2 -- 170.8] | [49.8 -- 702.8] | [7.0 -- 127.4] | [0.38 -- 486.8] | [7.0 -- 171.0] | [219.6 -- 471.7] | [34.0 -- 61.8] |
| OH | [20.4 -- 21.4] | [6.8 -- 6.9] | [51.7 -- 60.7] | [10.6 -- 10.9] | [41.3 -- 57.1] | [7.2 -- 8.2] | [14.1 -- 39.4] | [5.4 -- 6.8] | [9.9 -- 127.3] | [1.2 -- 14.1] |
| OK | [21.8 -- 30.6] | [6.3 -- 6.4] | [34.0 -- 56.0] | [8.0 -- 8.6] | [64.1 -- 89.7] | [11.4 -- 12.2] | [12.1 -- 74.3] | [4.2 -- 9.0] | [68.0 -- 81.9] | [11.8 -- 12.1] |
| OR | [5.8 -- 7.2] | [2.1 -- 2.2] | [10.9 -- 116.0] | [1.0 -- 10.7] | [45.9 -- 57.0] | [7.8 -- 9.3] | [3.9 -- 24.9] | [2.9 -- 5.0] | [11.9 -- 140.5] | [1.3 -- 12.8] |
| PA | [22.8 -- 24.5] | [8.2 -- 8.2] | [94.8 -- 106.1] | [19.3 -- 19.8] | [84.7 -- 98.9] | [16.2 -- 17.5] | [31.5 -- 56.4] | [10.2 -- 11.9] | [0.10 -- 268.5] | [1.3 -- 27.1] |
| RI | [23.1 -- 39.3] | [10.7 -- 11.1] | [79.9 -- 241.7] | [18.0 -- 23.2] | [59.7 -- 146.5] | [16.8 -- 19.3] | [25.2 -- 302.6] | [2.4 -- 19.7] | [0.83 -- 828.5] | [4.3 -- 78.2] |
| SC | [21.6 -- 25.3] | [6.9 -- 7.0] | [83.9 -- 93.1] | [14.8 -- 15.1] | [77.2 -- 93.4] | [8.1 -- 11.2] | [6.4 -- 71.1] | [2.3 -- 10.3] | [15.3 -- 197.6] | [2.2 -- 28.7] |
| SD | [29.7 -- 42.2] | [12.5 -- 12.8] | [30.3 -- 380.2] | [5.6 -- 77.0] | [50.1 -- 421.0] | [5.2 -- 49.0] | [54.1 -- 607.6] | [9.4 -- 95.7] | [228.9 -- 377.9] | [39.4 -- 43.6] |
| TN | [30.9 -- 35.5] | [7.9 -- 7.9] | [86.5 -- 94.1] | [13.9 -- 14.3] | [115.0 -- 154.4] | [14.0 -- 16.3] | [11.8 -- 79.0] | [3.3 -- 7.8] | [18.7 -- 302.4] | [1.3 -- 17.0] |
| TX | [29.1 -- 32.7] | [7.0 -- 7.0] | [68.3 -- 75.3] | [12.0 -- 12.3] | [142.7 -- 145.9] | [22.5 -- 22.6] | [18.8 -- 29.7] | [5.5 -- 6.2] | [30.2 -- 224.5] | [3.9 -- 12.0] |
| UT | [15.9 -- 19.7] | [4.7 -- 4.7] | [13.9 -- 169.1] | [3.5 -- 36.0] | [66.1 -- 90.5] | [13.2 -- 13.9] | [11.5 -- 68.9] | [4.9 -- 9.1] | [141.0 -- 403.8] | [18.0 -- 40.0] |
| VA | [12.5 -- 15.4] | [4.9 -- 5.0] | [45.0 -- 51.4] | [9.3 -- 9.7] | [73.9 -- 92.0] | [12.4 -- 14.1] | [11.6 -- 25.7] | [4.7 -- 5.7] | [4.5 -- 215.4] | [1.5 -- 18.6] |
| WA | [8.1 -- 11.1] | [3.5 -- 3.6] | [17.7 -- 50.9] | [5.3 -- 7.6] | [63.0 -- 77.1] | [12.9 -- 14.2] | [11.1 -- 26.6] | [3.8 -- 4.5] | [57.2 -- 133.9] | [9.1 -- 14.6] |
| WV | [11.8 -- 17.3] | [3.6 -- 3.7] | [13.9 -- 73.6] | [3.0 -- 7.0] | [0.23 -- 40.9] | [1.1 -- 14.2] | [14.2 -- 146.7] | [1.1 -- 15.5] | [17.3 -- 293.2] | [3.1 -- 54.5] |
| WI | [18.7 -- 22.7] | [7.4 -- 7.4] | [68.8 -- 108.0] | [17.2 -- 18.1] | [116.8 -- 139.7] | [20.1 -- 20.9] | [50.9 -- 83.1] | [9.7 -- 10.7] | [70.5 -- 226.3] | [14.6 -- 23.3] |

**Table S4**: Conservative 95% interval estimates of the age-adjusted YPLL rate attributable to COVID-19 and age-adjusted mortality rate attributable to COVID-19 for NH Whites, NH Blacks, Hispanics, NH Asians, and NH AIAN's in the U.S. and each examined state with respect to cumulative COVID-19 deaths according to data from the National Center for Health Statistics as of December 30, 2020. The upper reference age used to define YPLL is 75 years.
