## Supplementary material for "Racial and Ethnic Disparities in Years of Potential Life Lost Attributable to COVID-19 in the United States: An Analysis of 45 States and the District of Columbia": Table S5

| State | NH Black | | Hispanic | | NH Asian | | NH AIAN | |
|---|---|---|---|---|---|---|---|---|
| | YPLL | Mortality | YPLL | Mortality | YPLL | Mortality | YPLL | Mortality |
| US | [2.84 -- 2.97] | [1.91 -- 1.99] | [2.90 -- 3.02] | [1.95 -- 2.03] | [0.99 -- 1.17] | [0.83 -- 0.95] | [3.79 -- 5.06] | [1.82 -- 2.70] |
| AL | [2.79 -- 3.32] | [1.74 -- 1.81] | [2.63 -- 4.59] | [1.22 -- 1.95] | [0.20 -- 2.95] | [0.12 -- 1.57] | [0.24 -- 4.53] | [0.13 -- 2.19] |
| AZ | [1.86 -- 5.50] | [1.74 -- 2.87] | [4.41 -- 6.22] | [3.43 -- 3.79] | [0.73 -- 4.28] | [0.95 -- 2.10] | [14.79 -- 23.70] | [7.42 -- 8.97] |
| AR | [2.01 -- 2.64] | [1.42 -- 1.47] | [2.81 -- 4.20] | [1.78 -- 1.92] | [0.21 -- 2.86] | [0.17 -- 1.62] | [0.13 -- 2.42] | [0.19 -- 1.90] |
| CA | [3.31 -- 3.89] | [2.18 -- 2.29] | [6.16 -- 6.45] | [3.64 -- 3.69] | [1.25 -- 1.50] | [1.20 -- 1.25] | [1.32 -- 5.63] | [1.28 -- 2.36] |
| CO | [2.82 -- 6.50] | [2.04 -- 2.62] | [4.24 -- 6.41] | [2.76 -- 2.97] | [1.71 -- 6.55] | [1.60 -- 2.27] | [4.09 -- 22.40] | [1.26 -- 5.82] |
| CT | [3.55 -- 6.37] | [2.47 -- 2.73] | [2.79 -- 4.48] | [1.66 -- 1.90] | [0.71 -- 3.04] | [0.58 -- 1.12] | [0.01 -- 23.20] | [0.21 -- 7.44] |
| DE | [0.92 -- 4.39] | [1.55 -- 1.89] | [0.50 -- 6.21] | [0.63 -- 2.10] | [0.06 -- 2.01] | [0.17 -- 1.76] | [0.35 -- 13.67] | [0.79 -- 10.53] |
| DC | [3.91 -- 10.69] | [3.14 -- 3.65] | [8.36 -- 24.29] | [4.41 -- 5.97] | [0.67 -- 16.30] | [0.49 -- 4.64] | [1.22 -- 56.22] | [0.65 -- 10.26] |
| FL | [3.91 -- 4.30] | [2.80 -- 2.83] | [2.53 -- 2.82] | [2.21 -- 2.24] | [0.90 -- 1.75] | [0.92 -- 1.02] | [0.92 -- 10.31] | [0.21 -- 1.78] |
| GA | [2.66 -- 3.02] | [1.92 -- 1.96] | [3.32 -- 4.05] | [1.82 -- 1.94] | [0.58 -- 1.71] | [0.64 -- 0.81] | [0.34 -- 4.65] | [0.16 -- 2.20] |
| ID | [0.87 -- 21.78] | [0.87 -- 12.62] | [2.96 -- 6.96] | [2.46 -- 2.90] | [0.65 -- 12.42] | [0.29 -- 3.16] | [0.69 -- 14.50] | [0.40 -- 3.88] |
| IL | [4.13 -- 5.16] | [2.00 -- 2.10] | [5.60 -- 6.84] | [2.65 -- 2.79] | [1.20 -- 2.44] | [1.07 -- 1.26] | [0.49 -- 20.37] | [0.18 -- 5.28] |
| IN | [2.71 -- 3.67] | [1.89 -- 1.95] | [2.81 -- 3.80] | [1.51 -- 1.58] | [0.48 -- 2.04] | [0.81 -- 1.06] | [0.29 -- 11.12] | [0.16 -- 2.02] |
| IA | [2.61 -- 5.15] | [1.99 -- 2.20] | [3.19 -- 4.73] | [1.67 -- 1.82] | [0.66 -- 3.38] | [1.44 -- 1.83] | [2.02 -- 25.09] | [0.72 -- 8.00] |
| KS | [2.12 -- 7.58] | [1.82 -- 2.52] | [3.10 -- 6.58] | [2.26 -- 2.80] | [0.27 -- 7.74] | [0.42 -- 2.17] | [2.00 -- 31.90] | [1.04 -- 6.87] |
| KY | [2.08 -- 3.36] | [1.86 -- 2.02] | [3.65 -- 5.80] | [1.31 -- 2.07] | [0.37 -- 4.71] | [0.20 -- 1.79] | [0.02 -- 4.14] | [0.26 -- 4.07] |
| LA | [3.67 -- 4.38] | [2.20 -- 2.28] | [1.86 -- 3.96] | [1.10 -- 1.47] | [0.52 -- 2.24] | [0.35 -- 1.33] | [0.43 -- 7.60] | [0.16 -- 2.35] |
| MD | [3.19 -- 4.34] | [2.15 -- 2.24] | [7.20 -- 9.95] | [2.85 -- 3.21] | [0.77 -- 2.27] | [0.90 -- 1.09] | [0.47 -- 13.77] | [0.31 -- 4.03] |
| MA | [3.03 -- 4.79] | [1.87 -- 2.17] | [2.67 -- 3.98] | [1.59 -- 1.88] | [0.83 -- 2.11] | [0.86 -- 1.17] | [0.46 -- 30.33] | [0.23 -- 7.93] |
| MI | [5.77 -- 6.67] | [3.27 -- 3.48] | [3.09 -- 4.69] | [1.87 -- 2.69] | [0.34 -- 2.33] | [0.55 -- 1.43] | [0.71 -- 8.14] | [0.76 -- 4.44] |
| MN | [4.43 -- 6.67] | [2.40 -- 2.87] | [4.58 -- 8.64] | [2.01 -- 2.72] | [3.82 -- 6.05] | [1.75 -- 2.19] | [3.29 -- 13.73] | [1.48 -- 3.67] |
| MS | [2.71 -- 3.27] | [1.92 -- 1.98] | [1.09 -- 3.78] | [0.37 -- 1.42] | [0.57 -- 4.39] | [0.56 -- 2.32] | [17.97 -- 34.13] | [5.89 -- 9.46] |
| MO | [2.30 -- 3.07] | [1.67 -- 1.76] | [2.82 -- 3.97] | [1.25 -- 1.56] | [0.41 -- 2.31] | [0.83 -- 1.31] | [0.32 -- 5.05] | [0.18 -- 2.46] |
| MT | [3.26 -- 53.89] | [1.12 -- 26.66] | [1.55 -- 12.14] | [1.16 -- 3.52] | [0.91 -- 17.22] | [0.73 -- 6.41] | [11.14 -- 22.46] | [6.69 -- 7.46] |
| NE | [1.52 -- 7.38] | [1.51 -- 2.35] | [3.33 -- 7.45] | [2.40 -- 3.02] | [0.29 -- 6.31] | [0.19 -- 2.71] | [0.99 -- 20.04] | [0.59 -- 7.63] |
| NV | [2.49 -- 4.47] | [2.05 -- 2.21] | [3.42 -- 5.15] | [2.80 -- 2.93] | [1.25 -- 2.25] | [1.62 -- 1.73] | [1.47 -- 15.77] | [0.89 -- 3.94] |
| NH | [1.11 -- 54.19] | [1.04 -- 11.01] | [0.28 -- 21.60] | [0.42 -- 4.50] | [0.53 -- 25.86] | [0.37 -- 4.15] | [0.00 -- 57.70] | [0.00 -- 4.95] |
| NJ | [3.70 -- 3.99] | [2.20 -- 2.23] | [4.44 -- 4.76] | [2.36 -- 2.39] | [1.30 -- 1.78] | [1.13 -- 1.17] | [1.00 -- 10.92] | [0.25 -- 1.81] |
| NM | [0.48 -- 12.28] | [0.35 -- 3.06] | [1.43 -- 3.91] | [1.79 -- 2.04] | [0.12 -- 4.29] | [0.28 -- 3.24] | [11.83 -- 31.63] | [9.84 -- 11.31] |
| NY | [3.84 -- 5.76] | [2.77 -- 3.32] | [4.57 -- 6.49] | [2.94 -- 3.52] | [1.91 -- 3.80] | [1.55 -- 2.28] | [0.24 -- 29.90] | [0.29 -- 15.33] |
| NC | [3.16 -- 4.05] | [2.31 -- 2.42] | [6.57 -- 9.71] | [3.07 -- 3.53] | [0.69 -- 5.41] | [1.09 -- 1.82] | [0.71 -- 8.36] | [0.99 -- 2.61] |
| ND | [0.31 -- 15.37] | [0.45 -- 13.61] | [0.95 -- 21.94] | [0.48 -- 10.16] | [0.01 -- 14.99] | [0.51 -- 13.59] | [4.14 -- 14.58] | [2.51 -- 4.91] |
| OH | [2.43 -- 2.95] | [1.55 -- 1.59] | [1.95 -- 2.77] | [1.05 -- 1.20] | [0.66 -- 1.91] | [0.79 -- 0.99] | [0.47 -- 6.19] | [0.17 -- 2.06] |
| OK | [1.12 -- 2.53] | [1.25 -- 1.37] | [2.12 -- 4.06] | [1.78 -- 1.94] | [0.41 -- 3.34] | [0.66 -- 1.43] | [2.25 -- 3.69] | [1.83 -- 1.91] |
| OR | [1.55 -- 19.85] | [0.47 -- 5.04] | [6.57 -- 9.66] | [3.56 -- 4.39] | [0.57 -- 4.25] | [1.34 -- 2.33] | [1.74 -- 23.82] | [0.58 -- 6.03] |
| PA | [3.89 -- 4.61] | [2.34 -- 2.42] | [3.50 -- 4.29] | [1.97 -- 2.14] | [1.30 -- 2.45] | [1.24 -- 1.45] | [0.01 -- 11.72] | [0.16 -- 3.31] |
| RI | [2.08 -- 10.31] | [1.63 -- 2.17] | [1.55 -- 6.27] | [1.52 -- 1.81] | [0.67 -- 13.03] | [0.22 -- 1.84] | [0.01 -- 34.71] | [0.39 -- 7.28] |
| SC | [3.35 -- 4.25] | [2.13 -- 2.19] | [3.10 -- 4.25] | [1.16 -- 1.62] | [0.26 -- 3.22] | [0.32 -- 1.49] | [0.62 -- 8.89] | [0.32 -- 4.16] |
| SD | [0.73 -- 12.36] | [0.43 -- 6.14] | [1.22 -- 14.13] | [0.40 -- 3.90] | [1.36 -- 19.89] | [0.73 -- 7.58] | [5.61 -- 12.40] | [3.08 -- 3.48] |
| TN | [2.44 -- 3.02] | [1.75 -- 1.81] | [3.25 -- 4.96] | [1.77 -- 2.07] | [0.31 -- 2.54] | [0.42 -- 1.00] | [0.52 -- 9.63] | [0.16 -- 2.17] |
| TX | [2.10 -- 2.57] | [1.70 -- 1.76] | [4.38 -- 4.99] | [3.19 -- 3.24] | [0.58 -- 1.01] | [0.78 -- 0.89] | [0.93 -- 7.66] | [0.55 -- 1.72] |
| UT | [0.72 -- 10.38] | [0.75 -- 7.76] | [3.42 -- 5.58] | [2.80 -- 2.99] | [0.60 -- 4.23] | [1.03 -- 1.96] | [7.31 -- 24.85] | [3.81 -- 8.58] |
| VA | [2.94 -- 4.08] | [1.88 -- 1.98] | [4.84 -- 7.34] | [2.49 -- 2.87] | [0.75 -- 2.03] | [0.94 -- 1.16] | [0.27 -- 16.92] | [0.30 -- 3.81] |
| WA | [1.62 -- 6.11] | [1.45 -- 2.13] | [5.77 -- 9.34] | [3.56 -- 4.00] | [1.03 -- 3.21] | [1.03 -- 1.27] | [5.24 -- 16.17] | [2.52 -- 4.12] |
| WV | [0.86 -- 6.05] | [0.81 -- 1.95] | [0.01 -- 3.36] | [0.29 -- 3.93] | [0.84 -- 12.14] | [0.31 -- 4.28] | [1.04 -- 24.02] | [0.83 -- 15.22] |
| WI | [3.06 -- 5.71] | [2.32 -- 2.46] | [5.19 -- 7.38] | [2.71 -- 2.84] | [2.27 -- 4.39] | [1.31 -- 1.46] | [3.15 -- 11.94] | [1.96 -- 3.16] |

**Table S5**: Conservative 95% interval estimates of the age-adjusted COVID-19-attributable YPLL rate ratios and age-adjusted COVID-19-attributable mortality rate ratios for NH Blacks, Hispanics, NH Asians, and NH AIAN's relative to NH Whites in the U.S. and each examined state with respect to cumulative COVID-19 deaths according to data from the National Center for Health Statistics as of December 30, 2020. The upper reference age used to define YPLL is 75 years.
